# A surgical window of opportunity trial evaluating the effect of the PCSK9 inhibitor evolocumab on tumoral MHC-I expression and CD8^+^ infiltration in glioma

**DOI:** 10.1101/2024.06.19.24309192

**Authors:** Kirit Singh, Matthew W. Foster, Marlene J. Violette, Anna M. Corcoran, Kelly M. Hotchkiss, Chelsea O. Railton, Emily E. Blandford, Kathryn E. Blethen, William C McIntosh, Elizabeth L. Thomas, David M. Ashley, Annick Desjardins, Henry S. Friedman, Margaret O. Johnson, Allan Friedman, Stephen Keir, Evan D. Buckley, James E. Herndon, Roger E. McLendon, John H. Sampson, Evan Calabrese, Giselle Y. Lopez, Gerald A. Grant, Anoop P. Patel, Simon G. Gregory, Chuan-Yuan Li, Peter E. Fecci, Mustafa Khasraw

**Author notes:** These authors contributed equally to the manuscript (Peter E. Fecci, Mustafa Khasraw). **Corresponding Author.** Mustafa Khasraw, MD, The Preston Robert Tisch Brain Tumor Center, Duke University | Box 3624, Durham, NC 27710, |.

## Abstract

Many cancers evade immunosurveillance by downregulating surface major histocompatibility class (MHC)-I. Proprotein convertase subtilisin/kexin type 9 (PCSK9) promotes MHC-I degradation and is elevated in glioma. Evolocumab is a clinically approved PCSK9 inhibitor which restores MHC-I expression in pre-clinical cancer models. However, monoclonal antibodies have limited blood brain/tumor barrier penetrance (BBB/BTB). We conducted a window-of-opportunity trial, evaluating evolocumab’s BBB/BTB penetrance and biological effect (PesKE; NCT04937413). Patients with newly diagnosed or recurrent glioma undergoing a clinically indicated biopsy or resection were enrolled (n=32, M: 16, F: 16; control average age: 51.85, evolocumab: 53). Intervention participants (n=6) received a single subcutaneous evolocumab dose pre-procedure, of which 4 provided research tissue. No significant adverse events were observed. Evolocumab was detected in all analyzed intervention tissue, with an average tumor:blood ratio of 0.0222 (SD±0.0190), akin to other monoclonals. Evolocumab quantitation was 4.44x greater in contrast-enhancing (mean 0.0068 fmol/mcg (SD±0.001)) vs non-contrast enhancing cases (mean 0.0015 fmol/mcg (SD±0.0004)). Proteomic analysis found positive trends between evolocumab and MHC-I subtypes (HLA-A-C, E-G), with a significant positive correlation with HLA-H (R^2^=0.9584, p=0.021*). Tumor tissue with higher evolocumab titers demonstrated increased surface MHC-I and CD8^+^ T cell infiltration. Increased CD8^+^ *TNF*, *FASLG* and *GZMA* transcription was observed in high titer tissue compared to low titer tissue and untreated controls. Pre-resection evolocumab is well tolerated but exhibits BBB/BTB penetrance akin to other monoclonal antibodies. Increased tumoral evolocumab/PCSK9i may enhance tumoral MHC-I/effector CD8^+^ infiltration. Future work will explore combining evolocumab with BBB/BTB opening therapies like low-intensity focused ultrasound.

**One Sentence Summary:** We conducted a tissue-based study in glioma patients to evaluate if peripheral evolocumab enters tumors, enhances surface MHC-I, and boosts effector CD8^+^ T cell infiltration.

**Graphical Abstract:** 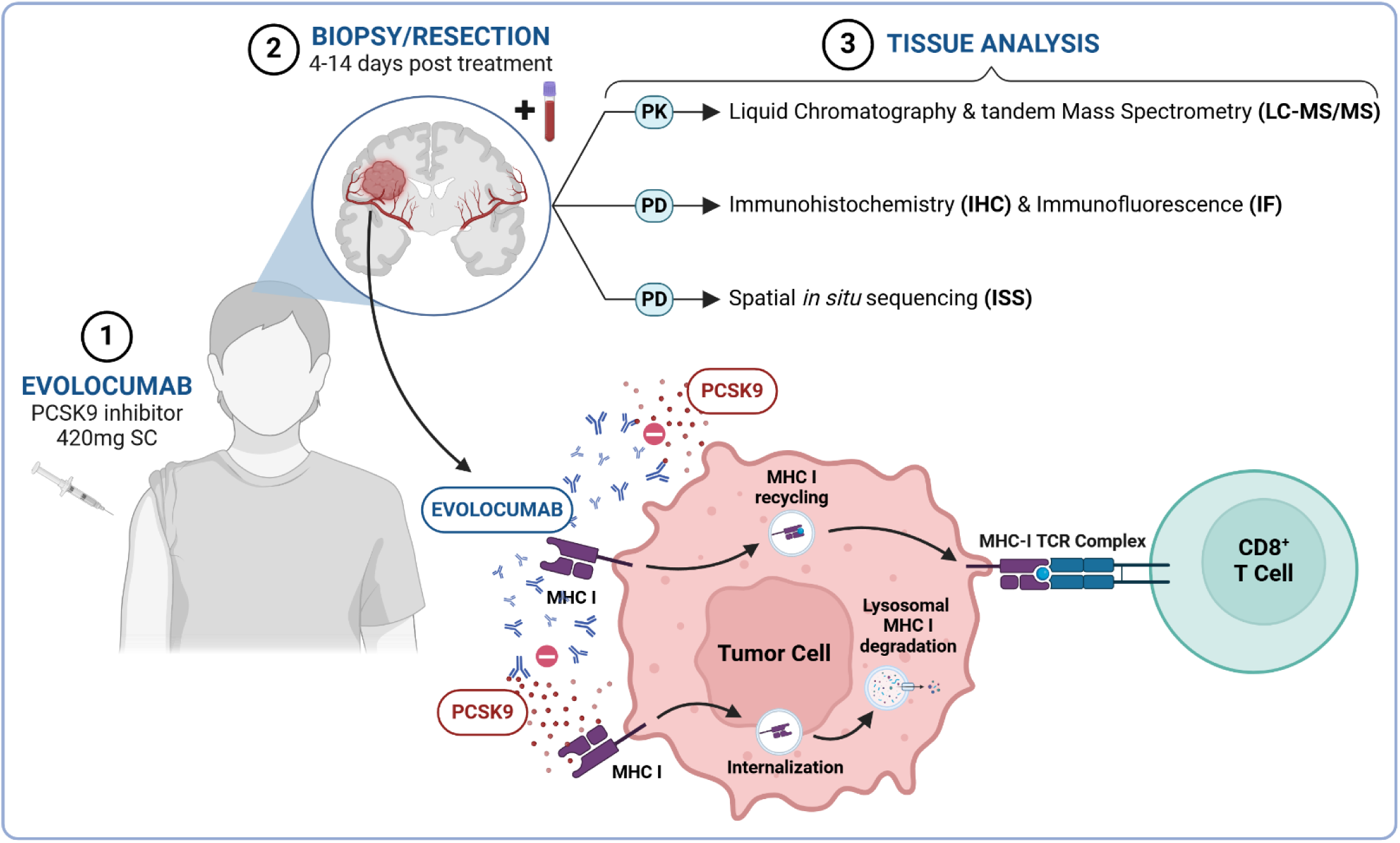

## INTRODUCTION

Tumoral evasion of the immune system is a key driver of resistance to immunotherapy. One such method of evasion is downregulation of surface major histocompatibility complex class I (MHC-I)^1^. Absence of MHC-I on the cell surface restricts neo-antigen presentation and conceals the tumor from cytotoxic CD8^+^ T cells. This phenomenon has been observed in many cancers, including primary brain malignancies like glioma^2^. Loss of MHC-I results in an immunologically cold tumor, with few tumor-infiltrating lymphocytes (TILs)^3^ and worsened clinical outcomes^4^. While recent findings by our group demonstrate that CD8^+^ killing can occur independently of MHC-I via the NKG2D-NKG2DL axis^5^, such killing depends on prior T cell priming via T cell receptor (TCR) activation. Restoring MHC-I expression may facilitate the priming of CD8^+^ T cells, thereby enabling them to kill the tumor^6,7^.

*Liu et al* recently established that regulators of cholesterol metabolism^8^, such as proprotein convertase subtilisin/kexin type 9 (PCSK9), can influence the recycling of MHC-I receptors between the plasma membrane and cytoplasm ^9^. PCSK9 was initially developed as a drug target for hypercholesteremia as it promotes the degradation of the low-density lipoprotein receptor (LDL-R)^10^. However, this effect has been found to translate across multiple surface receptors, including the very low-density lipoprotein receptor (VLDL-R), Apolipoprotein E receptor 2 (ApoER2) and cluster of differentiation 36 receptor (CD36)^10,11^. Similarly, *Liu et al* demonstrated that high levels of PCSK9 promote the internalization of MHC-I to intracellular lysosomes, where it is degraded^12^. Deletion/inhibition of PCSK9 blocks this pathway and increases tumoral expression of MHC-I, enhancing infiltration of cytotoxic CD8^+^ T cells (mechanism in **Fig. S1**). Analysis by Liu et al. of the Gene Expression across Normal and Tumor tissue (GENT) database gene expression database^13^ finds that PCSK9 mRNA expression negatively correlates with cytotoxic lymphocyte markers (CD8A) across multiple cancers, including esophageal cancer, cervical squamous cell carcinoma, endocervical adenocarcinoma and pancreatic adenocarcinoma^12^. When PCSK9 inhibition (PCSK9i) was combined with immune checkpoint blockade (ICB), such as anti-programmed-death 1 (αPD-1), enhanced anti-tumor efficacy was observed in in murine models of colon cancer and melanoma^12^. Interrogation of the pan-cancer cohort within the human protein atlas revealed that circulating plasma PCSK9 levels are increased in patients with glioma compared to other malignancies (**Fig. 1**)^14^. We have previously outlined the need for combination immunotherapies to overcome the multiple resistance pathways that are present in high-grade glioma^15^. Based on the findings by *Liu et al* and the elevated circulating levels of PCSK9 in glioma patients, we hypothesized that PCSK9i could reduce MHC-I degradation, thereby increasing tumoral MHC-I expression and enhancing the infiltration and activity of CD8^+^ lymphocytes. PCSK9i could then be considered as part of a future combination approach with T cell dependent therapies like ICB.

**Fig. 1.**
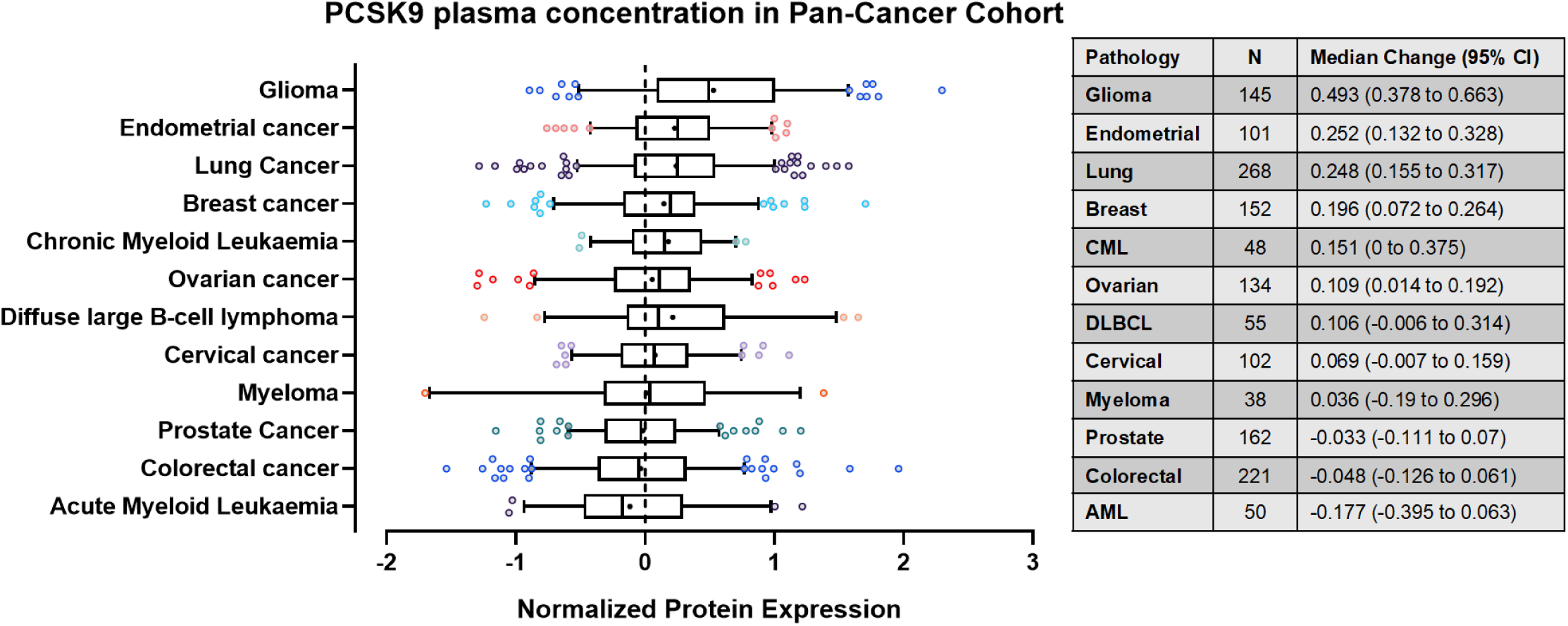
Relative plasma concentrations of PCSK9 in blood from patients with different cancer types. Box and whisker representations of circulating concentrations of PCSK9 protein in plasma across multiple cancer types. Circulating plasma PCSK9 protein levels are the most upregulated in glioma compared to the average concentration from samples across multiple. Assessed by proximity extension assay (PEA). Whiskers represent 5-95% range with outliers shown. Mean shown as **+**. Data from proteinatlas.org (https://www.proteinatlas.org/ENSG00000169174-PCSK9/disease).

Establishing this potential approach benefits from the commercial availability of PCSK9i therapies, which are approved by the Food and Drug Administration (FDA) for the management of hypercholesterolemia^16,17^. These include the monoclonal antibodies (mAbs) alirocumab^18^ and evolocumab^19^ as well as the small interfering RNA (siRNA) inclisiran^20^. We evaluated evolocumab as it had been used to demonstrate anti-tumor efficacy in combination with αPD-1 ICB by *Liu et al.* Evolocumab also possesses useful pharmacokinetic (PK) characteristics for combination therapy, having a well-established safety profile with the most common reported adverse event being mild injection-site reactions^16^. Further, at the maximum dose of 420 mg, evolocumab rapidly decreased unbound PCSK9 in the periphery within 4 hours of administration and fully suppressed PCSK9 levels up to 14 days following a single treatment^21^. Phase 3 clinical trials of evolocumab also found that no patients developed neutralizing antibodies to therapy over 2.2 years of follow-up^17^.

However, it is uncertain whether evolocumab readily cross the blood-brain/tumor barrier (BBB/BTB) as a mAb^22^. Although the BBB/BTB becomes disrupted in glioma via surgery, radiotherapy, or disease progression, the heterogenous uptake for antibody-based constructs^23^ is influenced by regions of the tumor that are shielded behind non-disrupted BBB/BTB and left untreated^24^. Despite this, evaluations of peripherally administered antibody therapy in the context of high-grade glioma (αPD-1) have demonstrated biological effect in the central nervous system (CNS) despite a cerebral-spinal fluid (CSF): serum ratio of just 0.009 ^25^. Further, evolocumab could theoretically associate with unbound PCSK9 in the periphery and prevent its downstream binding to tumor in the CNS, resulting in a similar pharmacodynamic (PD) effect.

Based on these data, we sought to determine whether evolocumab could affect the tumors of glioma patients and increase surface MHC-I, with resultant infiltration of cytotoxic CD8^+^ lymphocytes. To do so, we conducted a surgical window of opportunity (SWOOPP) study – The PCSK9i Inhibitor Evolocumab: A Surgical Trial of Pharmacodynamics and Kinetics Evaluation (PesKE, NCT04937413). The primary objective of this trial was to evaluate whether evolocumab could cross the BBB/BTB and be detectable in intracranial glioma following subcutaneous administration. Secondary objectives included determining the uptake ratio of evolocumab into the CNS by comparing levels in tumor vs blood and evaluating the downstream effects of PCSK9i on tumoral MHC-I abundance/expression and lipid metabolism. Exploratory analyses sought to evaluate changes in the characteristics of infiltrating immune cells between both groups. Assessments of long-term outcomes including Progression Free-Survival (PFS) and Overall Survival (OS) were not performed, as this was primarily a biomarker-focused study with limited patient enrollment. Safety monitoring for adverse events when using evolocumab in the context of glioma was also conducted throughout the study. We report summary findings from this trial and outline potential future combinatorial strategies using PCSK9 inhibition for intracranial malignancies.

## RESULTS

### Peripheral high-dose evolocumab is well tolerated in patients undergoing biopsy/resection of glioma

A total of 32 adult participants (≥ 18 years old) with newly diagnosed or recurrent glioma consented to enroll on study (16M, 16F) between October 2021 and June 2023. All enrolled patients had a clinical indication for surgical resection, debulking, or biopsy of their tumor. Participants were enrolled onto the treatment arm if it was feasible for them to receive evolocumab (420mg, subcutaneous) within 4-14 days before their planned procedure (trial design shown in **Fig. S2**). Of the 32 patients recruited to the study, 6 participants met inclusion criteria and consented to treatment with evolocumab (average age 53 (SD±19.88)), while 26 were assigned to the non-treatment group (average age 51.85 (SD±16.07). Full demographics and a breakdown of the diagnoses for both groups, including whether participants had newly diagnosed or recurrent glioma, is shown in **Table S1**). Tissue was taken for research if excess was available beyond that required for diagnostic purposes.

Of participants enrolled in the treatment arm, 5 patients proceeded to surgical resection/biopsy of their tumor. One treatment patient was not able to proceed to surgical resection. Four of the 5 treated patients who underwent resection had both post-treatment blood and tumor tissue available for analysis for the primary outcome. In the control arm, 17 participants had sufficient tissue and blood available for comparison (CONSORT^26^ flowchart showing tissue distribution shown in **Fig. 2**). Time from treatment with evolocumab to tumor resection was 4.4 days on average (SD±1.5). Over the course of the study, no grade 3-5 adverse events (AEs) or serious adverse events (SAEs) were recorded. The commonest were injection site reactions of the mild-moderate adverse events that were possibly, probably, or definitely related to evolocumab (n=2 (33%)). A summary of all adverse events recorded during this study is shown in **Table S2** with those possibly, probably, or definitely related to evolocumab shown in **Table S3**.

**Fig. 2.**
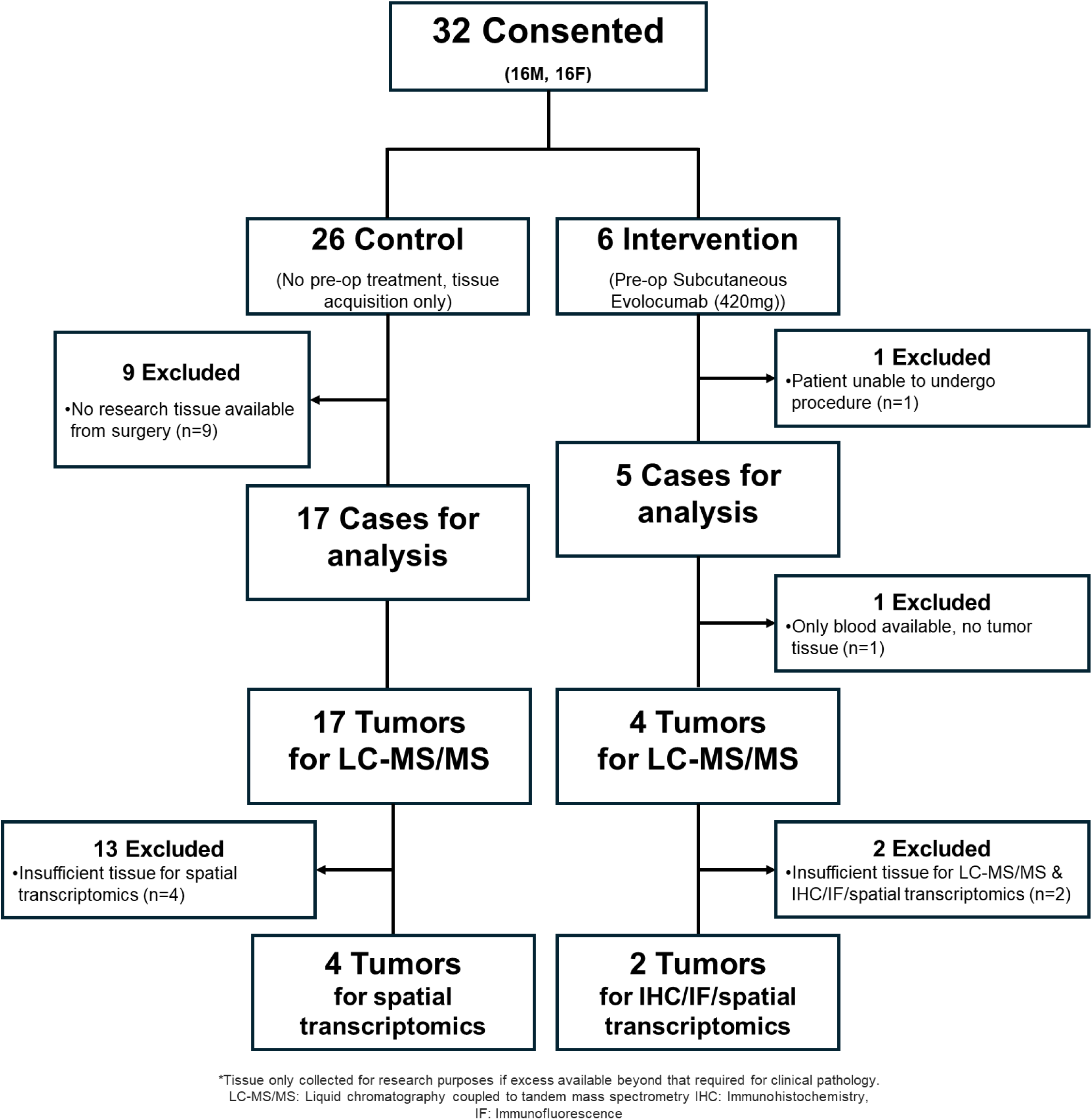
CONSORT flow diagram of enrolled participants.

### Peripherally administered evolocumab is detected in intracranial tumors, with enhanced titers in contrast-enhancing disease

To detect evolocumab accurately and specifically in tumor, we used liquid chromatography coupled to tandem mass spectrometry (LC-MS/MS). We selected three candidate “proteotypic” tryptic peptides after analysis of neat drug based on uniqueness versus reference human proteomes^27^ and validated these using drug after spiking into pooled human plasma. Evolocumab was quantified via a targeted LC-MS/MS assay that used stable isotope-labeled (SIL) internal standard peptides and an external calibration curve^28^. Using peptides which gave best figures-of-merit (i.e., lowest limits of detection and quantification), evolocumab was below limit of detection in blood prior to treatment but was quantified in all intervention patients following administration of drug, confirming specificity of detection (p=0.0286*, two-tailed Mann-Whitney U test, **Fig. 3A**). On evaluating tumor tissue, we were able to detect drug in all analyzed intervention cases, with minimal background signal observed in control tissue (intervention vs control arm; mean evolocumab quantitation of 0.0045 fmol/mcg (SD±0.0038) vs 1.4594 x 10^-5^ fmol/mcg (SD±2.6654 x 10^-5^) respectively, p=0.0015**, **Fig. 3B**). A positive, albeit non-significant, relationship was observed between evolocumab titers in blood and evolocumab titers in tumor (R^2^ =0.4020, p=0.2663, Pearson’s Correlation Coefficient, **Fig. 3C**). The average tumor: blood ratio of evolocumab was 0.0222 (SD±0.0190)), similar to other peripherally administered mAbs (αPD-1: 0.009^25^).

**Fig. 3.**
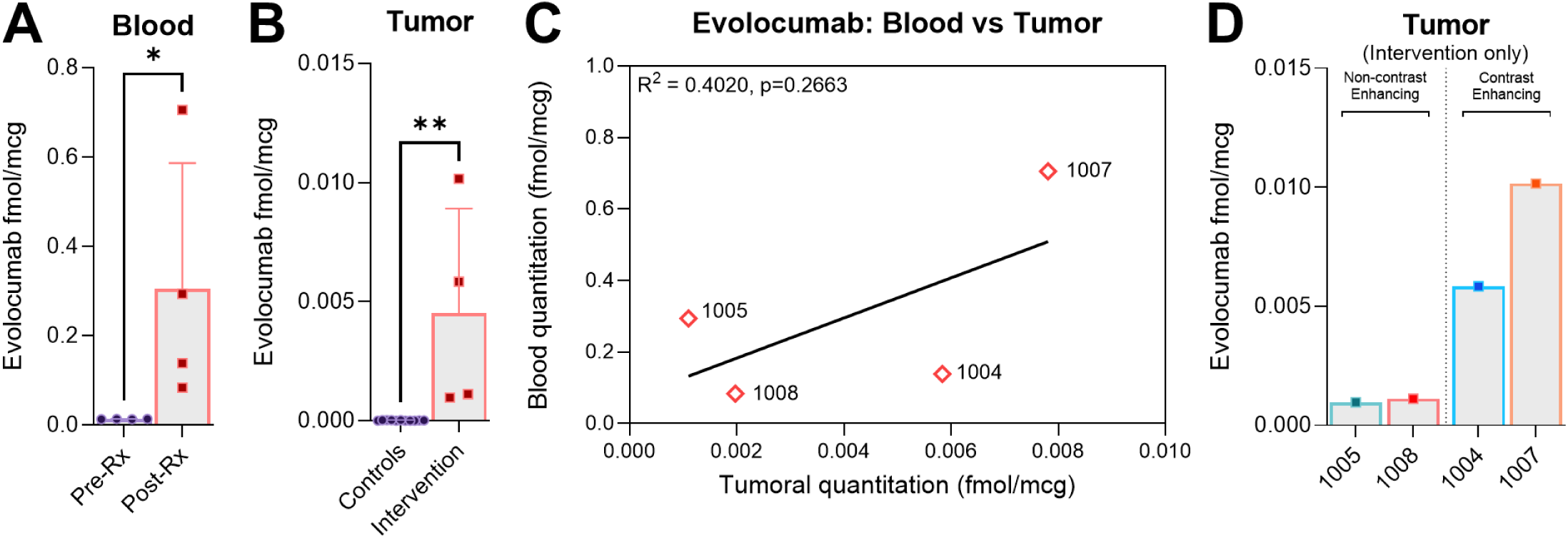
Peripherally administered evolocumab is detected in intracranial tumors, with enhanced uptake in contrast-enhancing disease. (A) Evolocumab levels in peripheral blood (1004: 0.1383 fmol/mcg, 1005: 0.2940 fmol/mcg, 1007: 0.7052 fmol/mcg, 1008: 0.0836 fmol/mcg). No evolocumab was detected in pre-treatment blood samples (p=0.0286*). Detection via most specific evolocumab detection peptide (GTMTTDPSTSTAYMELR) shown in analysis. Detection is normalized to stable internally labelled (SIL) peptide controls. Comparison via two-tailed Mann Whitney U test. Data presented as mean ± SD (B) Evolocumab levels in tumor compared to controls. Drug was detected in all intervention cases post subcutaneous administration, with negligible background signal detected in non-treatment controls (p=0.0015**, two two-tailed Mann Whitney U test). Data presented as mean ± SD (C) Scatterplot showing a positive but non-significant relationship between evolocumab titers in blood and evolocumab titers in tumor (R^2^ = 0.4020, p=0.2663, Pearson’s Correlation Coefficient). The average tumor: blood ratio of evolocumab was 0.0222 (SD±0.0190)). Average across individual detection peptides shown (sequences that are not part of the human proteome but identify drug: ASGYTLTSYGISWVR, GTMTTDPSTSTAYMELR and GYGMDVWGQGTTVTVSSASTK). Detection is normalized to stable internally labelled (SIL) peptide controls. (D) Evolocumab detection was higher in cases 1004 and 1007 (0.005838 fmol/mcg and 0.01016 fmol/mcg respectively), whereas detection was lower in cases 1005 and 1008 (0.000964 fmol/mcg and 0.001118 fmol/mcg respectively). The greatest uptake ratio was observed in the case with the highest degree of contrast-enhancement on MRI (ID: 1004, tumor:blood ratio of 0.0547, MRI shown in Fig. 4C).

Increased tumoral evolocumab titers were detected in intervention cases 1004 and 1007, with lower titers in cases 1005 and 1008 (**Fig. 3D**). Evaluation of pre-operative imaging and pathology for the two cases with lower evolocumab titers found that they were low grade non-contrast enhancing tumors (ID 1005: grade III Oligodendroglioma, ID 1008: grade II Astrocytoma, average evolocumab detection of 0.0015 fmol/mcg (SD±0.0004), magnetic-resonance imaging (MRI) sequences shown in **Fig. 4A-D**)). Conversely, cases with higher evolocumab titers were grade IV glioblastomas, with evidence of contrast enhancement on MRI imaging (IDs: 1004 & 1007, average evolocumab detection of 0.0068 fmol/mcg (SD±0.001), 4.44x compared to non-contrast cases). Though we had observed a positive relationship between levels of evolocumab in blood and levels of evolocumab in tumor, we noted that the greatest tumor: blood ratio was not in the case with the highest drug quantitation, but instead in the case with a more classical picture of ring enhancing contrast enhancement (ID: 1004, tumor: blood ratio of 0.0547, T1-post contrast MRI sequence shown in bottom left quadrant of **Fig. 4C**). In the other contrast-enhancing case (ID: 1007, T1-post contrast MRI sequence shown in bottom right quadrant of **Fig. 4D**), where enhancement was sparser, the tumor: blood ratio of 0.0117 was akin to non-enhancing lesions (ID: 1005 0.0073, ID: 1008 0.0151 tumor: blood ratios). We concluded that while subcutaneously administered evolocumab entered CNS tumor tissue, overall uptake was similar to other mAbs, with greater entry into tumors with greater BBB/BTB disruption.

**Fig. 4.**
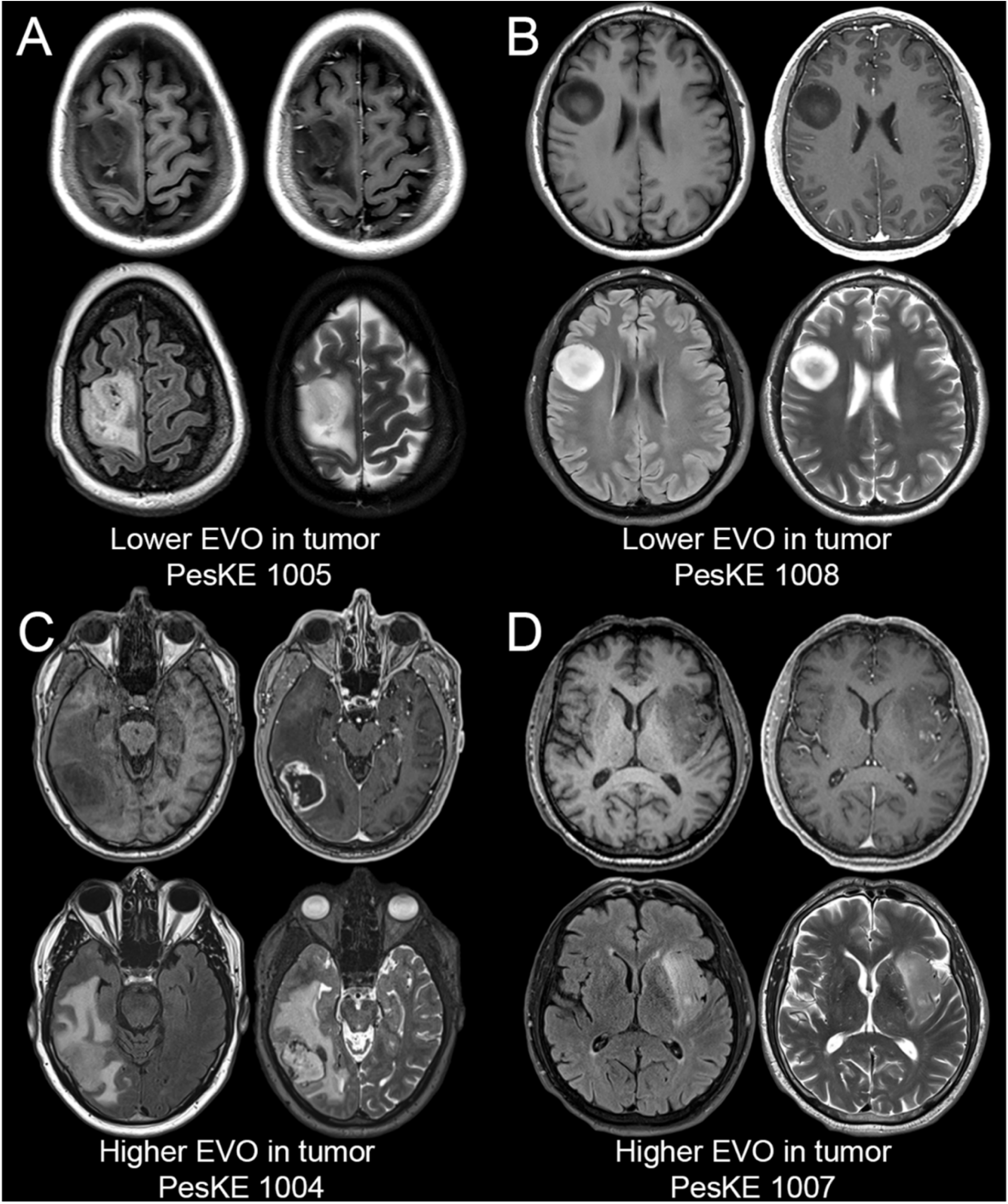
MRI sequences of tumors with lower and higher evolocumab (EVO) uptake. (A & B) Evaluation of pre-operative MR imaging from cases 1005 (grade III Oligodendroglioma) and 1008 (grade II Astrocytoma) with lower evolocumab titers reveals non-contrast-enhancing lesions on T1 post sequences (top row). (C & D) Conversely, evaluation of pre-operative MR imaging from cases 1004 and 1007 (both grade IV glioblastomas) with higher evolocumab titers reveals evidence of contrast-enhancement on T1 post sequences (bottom row). Representative pre-operative brain MRI images of analyzed tumors. For each panel A-D, axial images through the tumor are shown from four standard anatomic image series (clockwise from left: T1-weighted pre-contrast, T1-weighted post-contrast, T2-weighted, and T2-weighted FLAIR).

### Higher evolocumab uptake in tumor is associated with increased MHC-I and decreased Apolipoprotein E quantities within the tumor proteome

We next sought to evaluate the biological effect of evolocumab on tumor. We hypothesized that drug treatment would result in increased MHC-I levels in resected/biopsied tissue. To measure MHC-I levels, we initially anticipated using flow cytometry. However, limited tissue volumes from biopsy samples required us to use less tissue-intensive methods, and we therefore performed non-targeted tumor proteome analysis via LC-MS/MS on samples used for initial drug quantitation. First, we began by evaluating if non-targeted LC-MS/MS could appropriately determine high or low MHC-I expression on glioma cells. To do so, we analyzed immortalized glioma lines that have been reported in the literature to express either high levels of MHC-I (U343MG, MHC-I^Hi^) or lower levels of MHC-I (U87MG, MHC-I^Lo^)^29^. We confirmed the relative expression of MHC-I (HLA-ABC) via flow cytometric analysis (**Fig. 5A**) before analyzing the same cell lines via LC-MS/MS. Similar trends in relative levels of HLA-A, -B and -C were quantified in the cell proteomes from tryptic digests by non-targeted proteomics, with higher levels observed in the MHC-I^Hi^ U343MG versus MHC-I^Lo^ U87MG line (**Fig. 5B**). These data validated the use of LC-MS/MS for quantification of relative MHC-I.

**Fig. 5.**
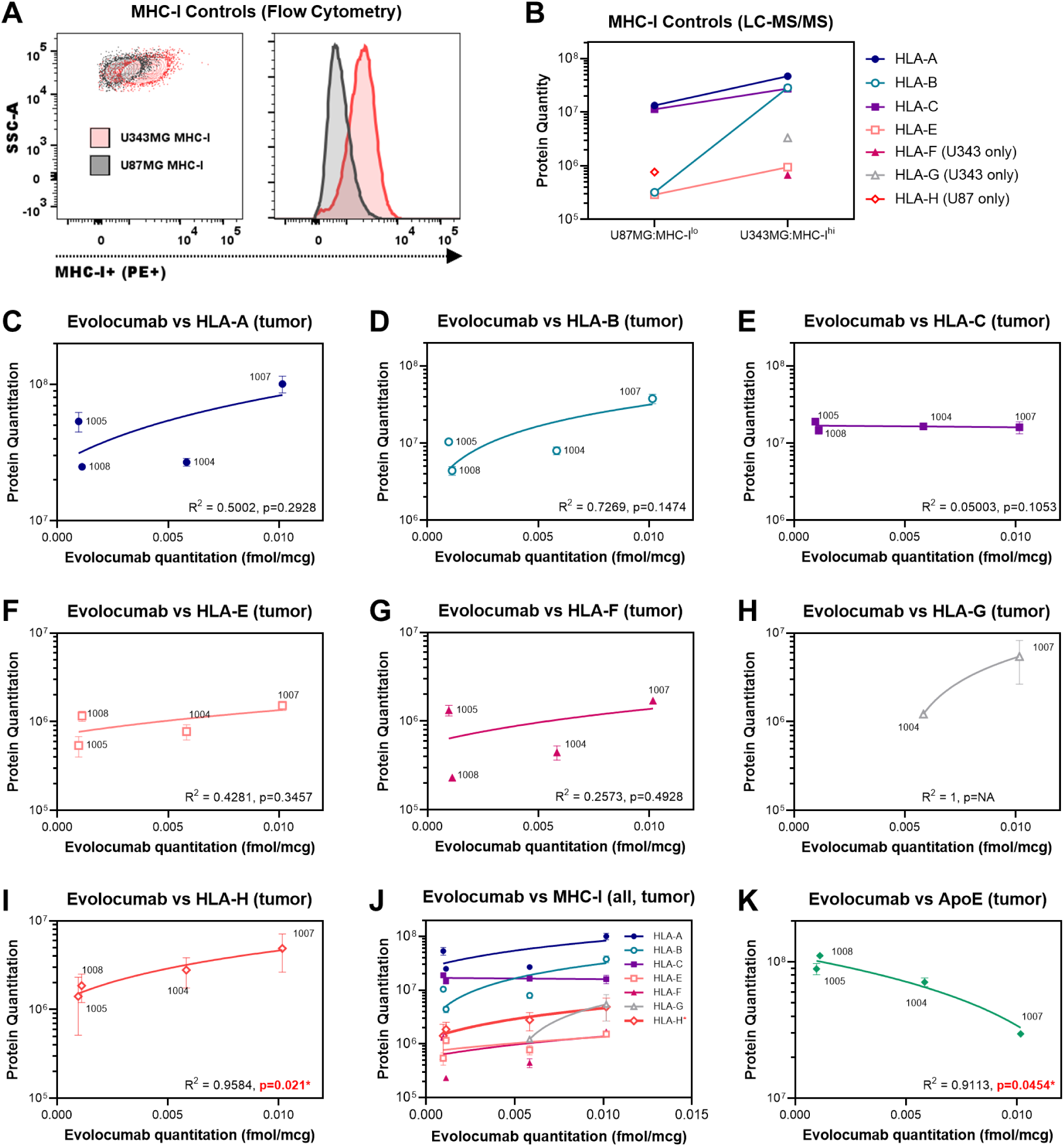
Higher evolocumab titers in tumor is associated with increased MHC-I and decreased Apolipoprotein E quantities within the tumor proteome. (A) Flow cytometric analysis of immortalized human glioma lines thought to express higher (U343MG, red) or lower (U87MG, black) levels of MHC-I. Staining for HLA-ABC confirms that U343MG is MHC-I^Hi^, while U87MG is MHC-I^Lo^. (B) Non-targeted LC/MS-MS quantitation of the tumor proteome reveals that U343MG has higher overall protein quantities of HLA-A, -B, -C and -E, compared to U87MG (HLA-A: U343MG vs U87MG 4.67 x 10^7^ vs 1.33 x 10^7^, HLA-B: 2.85 x 10^7^ vs 3.22 x 10^5^, HLA-C: 2.75 x 10^7^ vs 1.13 x 10^7^, HLA-E: 9.43 x 10^5^ vs 2.91 x 10^5^). Non-targeted proteome analysis yielded incomplete data for HLA-F, HLA-G and HLA-H (data included). These findings were in keeping with our flow cytometric analysis in (A) and provides validation for our detection method for MHC-I subtypes in tumors. (C) A positive but non-significant correlation was observed between intratumoral evolocumab and HLA-A protein levels (R^2^=0.5002, p=0.2928, n=4). (D) Similar trends were observed between intratumoral evolocumab and HLA-B protein levels (R^2^=0.7269, p=0.1474, n=4). (E) No change in HLA-C levels were observed in response to intratumoral evolocumab (R^2^=0.05003, p=0.1053, n=4). (F-H) Positive but non-significant correlative trends were also observed for HLA-E (R^2^=0.4281, p=0.3457, n=4), HLA-F (R^2^=0.2573, p=0.4928, n=4) and HLA-G (R^2^=1, p=NA, n=2, incomplete non-targeted proteome analysis yielded no data for ID: 1005 or ID: 1008 via LC-MS/MS). (I) A significant positive correlation was observed between intratumoral evolocumab and HLA-H protein levels (R^2^=0.9584, p=0.021*, n=4). (J) Trends across all MHC-I subtypes in relation to intratumoral evolocumab shown. (K) Evaluation of Apolipoprotein E levels across intervention samples found a significant negative trend between ApoE and intratumoral evolocumab (R^2^=0.9113, p=0.0454*). Data presented as mean ± SD unless otherwise specified. Correlation analyses were performed using Pearson’s Correlation Coefficient (PCC). Trendlines shown reflect simple linear regression.

Analysis of all MHC-I subtypes via LC-MS/MS revealed positive trends between the amount of Evolocumab in tumor and quantified protein levels of HLA-A (**Fig. 5C**), HLA-B (**Fig. 5D**), HLA-E (**Fig. 5F**), HLA-F (**Fig. 5G**) and HLA-G (**Fig. 5H**, non-targeted proteomic data only available for 2/4 cases), although these correlations were non-significant. We did, however, observe a significant positive correlation between Evolocumab and HLA-H in tumor (R^2^=0.9584, p=0.021*, Pearson’s Correlation Coefficient, **Fig. 5I**). Overall, broadly similar trends across MHC-I subtypes and evolocumab levels were observed, with the exception of HLA-C which remained unchanged (HLA-C shown in **Fig. 5E**, trends across all subtypes shown in **Fig. 5J**).

We next evaluated if increased evolocumab levels in tumor also resulted in changes in lipid pathways within the brain. On reviewing the non-targeted proteomic analysis for relevant markers, we first considered PCSK9 itself. However, LC/MS-MS quantitation of PCSK9 was incomplete for intervention samples, precluding analysis. We instead reviewed other regulators of lipid metabolism with the CNS. Noting that PCSK9 has been implicated in the degradation of Apolipoprotein E (ApoE) receptors, and that ApoE is the major apolipoprotein regulating CNS lipid metabolism^30^, we next analyzed relative ApoE abundances. Non-targeted proteomic data was available for ApoE levels in all intervention cases. We hypothesized that evolocumab would prevent degradation of ApoE receptors^10^ and therefore decrease ApoE protein levels. Indeed, we observed a significant negative correlation between ApoE protein levels and intratumoral evolocumab, indicating greater pharmacodynamic effect with greater tumoral titers of drug (R^2^=0.9113, p=0.0454*, **Fig. 5K**).

### Increased cytotoxic CD8^+^ T cell infiltration and MHC-I cell surface expression is observed near proliferating tumor cells in tissue with higher evolocumab titers

Finally, we sought to establish whether the increased MHC-I abundance in the proteome reflected increased expression on tumor. To do so, we performed parallel spatial transcriptomics (ST), immunofluorescence (IF), and immunohistochemistry (IHC) in tumor tissue with either lower (ID: 1008) or higher (ID: 1007) evolocumab uptake. Using ST and H&E staining, we classified various cell subtypes using canonical genes and identified regions of tumor, including proliferating (Ki-67^Hi31^) malignant cells (exemplar ID: 1007 in **Fig. 6A**, niches in **Fig. 6B**, Ki-67^Hi^ tumor cells and CD8^+^ lymphocytes shown in **Fig. 6C**, clustering analysis and identification in **Fig. S3- 4**, H&E in **Fig. S5**).

**Fig. 6.**
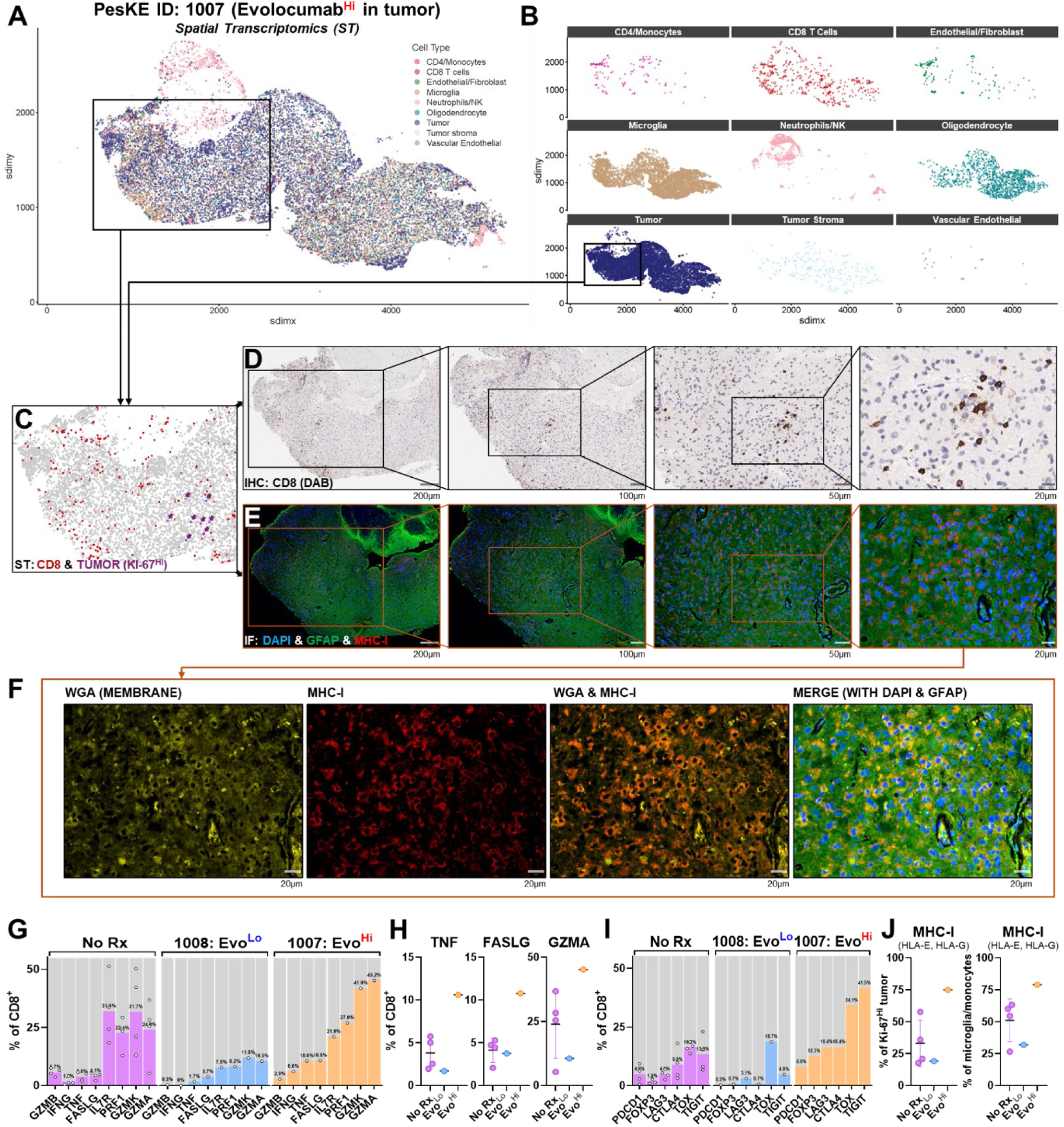
Increased cytotoxic CD8^+^ T cell infiltration and MHC-I cell surface expression is observed near proliferating tumor cells in tissue with higher evolocumab titers. (A & B) Paired spatial transcriptomics via Xenium platform and individual niches shown. Boxed region indicates areas of tumor as assessed by H&E and transcriptomic analysis. (C) Area of interest shown with Ki-67^Hi^ (i.e., proliferating) tumor highlighted in purple (with arrows) and CD8^+^ T cells highlighted in red, as identified via spatial transcriptomics. Overall tumor is shown in background (gray). (D-F) Paired Immunohistochemistry (IHC) and Immunofluorescence (IF) from tissue samples from a case with higher evolocumab titers adjacent to region of Ki-67^Hi^ tumor (ID: 1007, grade IV glioblastoma). High levels of parenchymal CD8^+^ infiltration within the parenchyma adjacent to proliferating tumor cells are observed on IHC staining. IF staining of GFAP (green), DAPI (nuclei, blue) and MHC-I (red) shows increased MHC-I expression in regions which co-localized with areas of CD8^+^ infiltrate. Staining with WGA (yellow) demonstrates MHC-I co-localized with cell membranes. (G) Transcriptomic comparison of the fraction of CD8^+^ cells expressing markers of cytotoxic activity (*GZMB, IFNG, TNF, FASLG, PRF1, GZMK, GZMA*) between untreated controls, and tumor tissue with low (ID: 1008) or high (ID: 1007) evolocumab titers. (H) Evaluation of most differentially expressed markers reveals that *TNF*, *FASLG* and *GZMA* were most increased in tumor tissue with high (ID: 1007) evolocumab titers (evolocumab high vs non-treated % CD8^+^: *TNF* 2.78x, *FASLG* 2.62x, *GZMA* 1.89x). (I) Transcriptomic comparison of the fraction of CD8^+^ cells expressing markers of T cell exhaustion (*PDCD1, FOXP3, LAG3, CTLA4, TOX, TIGIT*) between untreated controls, and tumor tissue with low (ID: 1008) or high (ID: 1007) evolocumab titers. (J) Transcriptomic comparison of levels of MHC-I subtypes HLA-E and HLA-G (available in panel) in (left) Ki-67^Hi^ tumor cells and (right) in microglia/monocytes. Comparison between untreated controls, and tumor tissue with low (ID: 1008) or high (ID: 1007) evolocumab titers. Data presented as mean ± SD unless otherwise specified.

Interestingly, ST revealed transcriptional evidence of focal parenchymal aggregates of CD8^+^ lymphocytes throughout tumor tissue with high evolocumab titers, including adjacent to proliferating tumor cells (**Fig. 6C**). Unblinded pathologist review also identified parenchymal aggregates of CD8^+^ cells in IHC staining of the same tissue, noting that this behavior differed from the typical perivascular pattern seen in glioma. Curious as to whether this increased CD8^+^ infiltration correlated with MHC-I changes, we conducted a focused IF analysis of the area where ST/IHC labelled CD8^+^ aggregates were adjacent to proliferating tumor cells. In these regions, we observed high MHC-I expression via IF, which notably overlapped with the membrane marker Wheat Germ Agglutinin (WGA, focused CD8^+^ IHC sequence in **Fig. 6D**, paired focused IF of DAPI, GFAP, MHC-I and WGA in **Fig. 6E & 6F**). Similar trends were not seen in tumor tissue with lower evolocumab titers, where sparse CD8^+^ infiltration on ST/IHC and reduced co- membranous MHC-I expression was seen (paired ST, CD8^+^ IHC and MHC-I IF in **Fig. S6**, H&E in **Fig. S7**).

Having observed enhanced CD8^+^ infiltration adjacent to proliferating tumor cells, we next considered the functional status of these lymphocytes. CD8^+^ activity across the whole sample was compared between untreated controls, low evolocumab titer tissue and high evolocumab titer tissue using ST. Comparisons were made via transcriptomic analysis using canonical markers of effector T cell activity (*GZMB, IFNG, TNF, FASLG, PRF1, GZMK, GZMA*) (**Fig. 6G**). For tissue with high evolocumab titers, transcriptional levels of the cytotoxic genes *TNF*, *FASLG* and *GZMA* were increased above levels in both untreated controls and tissue with low evolocumab titers (evolocumab high vs non-treated % CD8^+^: *TNF* 2.78x, *FASLG* 2.62x, *GZMA* 1.89x, **Fig. 6H**). We also evaluated the transcription of T cell exhaustion genes (*PDCD1, FOXP3, LAG3, CTLA4, TOX, TIGIT*) and observed increases of these markers in high evolocumab titer tissue compared to untreated and low titer samples (**Fig. 6I**). Further analysis of MHC-I transcription (HLA-E, HLA-G subtypes available in xenium panel) aligned with our IF findings. Interestingly, MHC-I transcription was also increased on antigen presenting cells (APCs, microglia/monocytes) in high evolocumab titer tissue compared to low titer tumor and untreated controls (**Fig. 6J**). Together, these data suggested increased tumoral/APC expression of MHC-I in tissue with a high evolocumab level. This increased MHC-I expression was accompanied by parenchymal CD8^+^ infiltration and evidence of effector T cell activity.

## DISCUSSION

Immunologically cold tumors like high-grade gliomas have proven resistant to immunotherapies such as immune checkpoint blockade ^32^. Loss of MHC-I has long been considered to be a key factor in resistance to immunotherapy, although recent findings by our group demonstrate that CD8^+^ killing can occur independently of MHC-I via the NKG2D-NKG2DL axis ^5^. However, such killing depends on prior TCR activation and restoring MHC-I expression may also facilitate this priming of CD8^+^ anti-tumor activity.

We performed a small-scale tissue-based trial to assess both the PK and PD of evolocumab in glioma – a drug that has been shown to increase MHC-I expression and potentiate ICB in other cancers

*in vivo*. We report that evolocumab appears to exhibit limited BBB/BTB penetrance (average tumor:blood ratio of 0.0222 (SD±0.0190)), in keeping with other mAbs that have not demonstrated efficacy against glioblastoma^25,32^. However, we did observe PD changes that correlated with levels of intratumoral evolocumab. Across the majority of MHC-I subtypes, we observed positive relationships between intratumoral drug levels and proteomic quantitation of HLAs, with a significant positive correlation between intratumoral evolocumab and HLA-H. Interestingly, while HLA-H was initially characterized as a non-functional pseudogene^33^, recent findings suggest evidence of transcriptional activity^34^ including roles in mobilizing other HLA subtypes to the cell surface^35^. Subsequent analysis via paired spatial ST/IHC/IF found that tumor tissue with higher evolocumab titers demonstrated higher levels of membranous MHC-I expression. These regions of high membranous MHC-I co-localized with parenchymal CD8^+^ infiltration, an uncharacteristic finding in higher-grade gliomas^36,37^. Additionally, we observed increased MHC-I transcription on APCs in tumor with higher evolocumab titers along with evidence of corresponding CD8^+^ effector activity. Increased expression of MHC-I on both tumor and resident APCs is known to drive CD8^+^ activation and subsequent proliferation, which may account for the observed increase in parenchymal CD8^+^ population^38^. Indeed, we find that transcription of markers of CD8^+^ T cell activity (*TNF, FASLG, GZMA*) and subsequent exhaustion (*PDCD1, FOXP3, LAG3, CTLA4, TOX, TIGIT*) are increased in tumor tissue with high evolocumab titers.

Further correlative evidence of increased biological activity was demonstrated by the significant inverse relationship between intratumoral evolocumab and levels of ApoE, a key regulator of CNS lipid metabolism (ApoE^30^). As mentioned previously, evolocumab has been shown to prevent loss of the ApoER2 receptor^10^, which internalizes and promotes the degradation of ApoE. This observed effect of evolocumab reducing ApoE levels in the CNS may also be of value, given that ApoE is over-expressed in high and low-grade gliomas^39^. In particular, ApoE induces immunosuppressant signaling in the CNS^40^ including the polarization of macrophages from a pro-inflammatory M1 to an anti-inflammatory M2 phenotype^41,42^. Given that immunosuppressive tumor associated macrophages (TAMs) are abundant in high-grade gliomas^43^, this effect would likely be beneficial in promoting a more efficacious anti-tumor immune response. Therapeutic targeting of ApoE and its isoforms in the CNS may also be of value in other disease areas, including neurodegenerative diseases, such as Alzheimer’s^44,45^.

Despite these promising findings, our study is limited by the small sample size of the treatment arm, with analysis of relationships between intratumoral evolocumab and PD parameters such as MHC-I subtypes/ApoE limited to just 4 cases. This limitation was likely driven by the treatment course of a single dose of evolocumab, potentially reducing patient interest in the study. Future tissue-based studies will benefit by offering sustained therapeutic intervention after tissue acquisition to facilitate patient enrollment. Another potential confounder for interpreting our findings include the potential of peripheral blood contamination in surgically acquired tissue samples. This is particularly problematic when attempting to detect low levels of drug in small amounts of tissue (as will likely be the case in many SWOOPP studies conducted in the CNS). While there are processes to minimize cross contamination, ranging from the simple (e.g., rinsing tissue with saline to remove excess blood, as was performed in this study) to complex (normalization to quantitated hemoglobin protein levels in tissue and blood), there is little validation regarding the relative efficacy of these. Ultimately, standardized processes to assess drug quantitation (PK) and biological effect (PD) are required. This will require harmonization of approaches to remove confounding drug signals from peripheral blood and agreed LC-MS/MS protocols for quantitation of protein levels in tissue.

Nevertheless, small-scale Window of Opportunity studies afford us the ability to quickly assess the impact of treatment. Given the suggestion of increased PD effect with greater PCSK9i in tumor, but PK characteristics of evolocumab that are similar to other mAbs, we could rationalize early modification of our treatment approach based on these initial findings. Modifications might include considering combination alongside modalities that increase the uptake of larger constructs into the brain such as low-intensity focused ultrasound (LIFU) via transient BBB/BTB opening^46,47^ or lipid nanoparticle encapsulation of PCSK9 siRNA^48^ to aid BBB/BTB penetration. Notably though, we observed increased transcription of markers associated with both cytotoxicity and exhaustion in CD8^+^ T cells which entered tumor tissue with high evolocumab titers. A combination strategy leveraging BBB/BTB opening to facilitate the entry of therapies that upregulate MHC-I and prevent exhaustion may further enhance intra-tumoral CD8^+^ T cell activity.

In conclusion, pre-operative high dose evolocumab in patients undergoing resection or biopsy for glioma appears to be well tolerated. However, the uptake of evolocumab across the BBB/BTB is in keeping with other antibody therapies, with the highest uptake observed in cases with the greatest degree of contrast-enhancement. Despite this, we find suggestions of mechanistic effect of PCSK9i in the context of high-grade glioma. Increases across the majority of MHC-I subtypes and decreases in ApoE levels were observed as intratumoral evolocumab levels increased. In tumor tissue with high evolocumab titers, increased tumoral/APC MHC-I expression and CD8^+^ T cell infiltration was observed. Future work will explore combination strategies that can enhance evolocumab uptake into high-grade gliomas, including low-intensity focused ultrasound and the addition of ICB therapies to prevent infiltrating CD8^+^ exhaustion.

## MATERIALS AND METHODS

### Study oversight and design

PesKE was a non-randomized, open-label, single-center surgical window of opportunity study. This study recruited adult patients (≥ 18 years old) with newly diagnosed or recurrent glioma and who had a clinical indication for either gross macroscopic resection, debulking, or biopsy of their disease. Participants were enrolled if pre-operative imaging indicated a sufficient tumor size that would allow for collection of specimens for the required analyses. A full list of final eligibility criteria is included in **Table S4**.

Once enrolled, participants on the treatment arm received 420mg of Evolocumab (administered subcutaneously) 4-14 days prior to their surgical procedure. Participants were assigned to the treatment arm if it was logistically feasible for them to receive subcutaneous evolocumab prior to their planned procedure (i.e., patients could attend for pre-operative administration of drug within the study’s timeframe of 4-14 days prior to procedure). Participants assigned to the control arm received no treatment prior to surgery. All participants enrolled in the study were assigned to have an intra-operative blood draw at the time of tissue collection. Treatment patients also underwent a blood draw prior to evolocumab administration. Tumor tissue for research purposes was only taken after sufficient tissue for clinical diagnostic purposes was collected. Participants on the treatment arm were then followed up for two weeks following surgery or biopsy and monitored for adverse events as well as routine post-operative assessments of physical and neurological status as well as standard laboratory evaluations. An overview of the study design is shown in **Fig. S2.**

The study received approval by the institutional review board at the Duke Cancer Institute (DCI, Duke IRB# Pro000108375) and was conducted at the same center. The study was registered on ClinicalTrials.gov (NCT04937413) on June 16th, 2021, and the first patient was enrolled on October 18th, 2021. The study was completed on June 21st, 2024. Study monitoring was performed both internally by the Principal Investigator (PI) and institutionally by the DCI. Audits of compliance with the protocol and principles of Good Clinical Practice were routinely performed by the Duke Office of Audit, Risk and Compliance (OARC). Data and Safety Monitoring were performed in accordance with the DCI Data and Safety Monitoring Plan. A full copy of the protocol is available on ClinicalTrials.gov.

### Tissue handling

Following surgery, samples were received in the surgical pathology laboratory and examined by the diagnostic staff. Areas suspicious for tumor were examined by frozen section by a neuropathologist. Once the diagnosis of tumor was established, tissue in excess of that required for an adequate and accurate diagnosis were collected and processed by members of the technical staff of the Duke Brain Tumor Biorepository (BTBR), a College of American Pathologists certified biorepository. Tissues were snap-frozen in liquid nitrogen either plain or embedded into Optimal Cutting Temperature (OCT) compound. If tissue samples were excessively bloody, they were rinsed with saline prior to freezing. Time as well as sample dimensions and weights were recorded at the point of freezing. At the time of surgery, whole blood was collected in tubes containing Ethylenediaminetetraacetic acid (EDTA) and placed on dry ice for transfer to storage. After freezing, samples were stored in liquid nitrogen and transferred to -80 °C prior to proteomic analysis. Sample collection details were maintained in the BTBR’s Nautilus laboratory information management system (LIS), with clinical details maintained in REDCAP. After samples were checked into the BTBR LIS, they were then distributed to research teams for downstream analysis.

### Cell Lines & Flow Cytometry

Cell lines (U87MG, U343MG) were purchased from the American Type Culture Collection (ATCC). Genetic testing and PCR evaluation performed via IDEXX bioanalytics (U87MG: # 124718-2023, U343MG: # 124717-2023). All lines tested negative for mycoplasma per IDEXX screening. Flow Cytometry staining of MHC-I was performed using 20uL/million cells of HLA-ABC (PE), BD #560964. Live/Dead staining performed using L/D Aqua (Invitrogen, L34966).

### Immunohistochemistry & Immunofluorescence

IHC and IF sectioning, staining, and imaging were performed at an external vendor (HistoWiz Inc). Staining was performed via a Leica bond automated staining platform, Akoya imager, and a proprietary processing, embedding, and grossing workflow. IHC stains were performed for CD45^+^ (1:100, Abcam #ab40763) and CD8^+^ (1:200, Leica #NCL-L-CD8-4B11). Triplex IF co-stained for Wheat Germ Agglutinin (WGA, 1:100, Invitrogen #W11261), MHC-I (1:3000, Cell Signaling #88274) and Glial Fibrillary Acidic Protein (GFAP, 1:6000, Novus #NB300-141) with 4’,6-diamidino-2-phenylindole (DAPI) counterstain.

### Mass Spectroscopy

For targeted assay development, 30 µg of neat evolocumab, or 30 or 100 pmol evolocumab in 20 µL of pooled human plasma (Golden West Diagnostics) were processed as previously described^28^. A complete description of targeted assay development is included within the supplementary materials.

For targeted quantification of evolocumab in whole blood, 20 µL of EDTA-blood was spiked with 2 pmol of JPT SpikeTides TQL peptides with the sequences ASGYTLTSYGISWVR, GTMTTDPSTSTAYMELR and GYGMDVWGQGTTVTVSSASTK (and 10 pmol of evolocumab in positive control samples) followed by deoxycholate-assisted trypsin digestion and microflow LC-MS/MS with solvent divert as described previously^28^. Briefly, deoxycholate-assisted trypsin digestion was used above except that 500 µg of TPCK-trypsin was used, and reactions were quenched with 2% TFA. An external calibration curve was generated using evolocumab in whole blood from a control subject, and quantification to standard curve was performed using Skyline. Blood levels (normalized to volume) were further normalized to microgram protein (fmol/µg) based on an estimated protein concentration in blood of 250 µg/μL.

Fresh frozen tissue either directly frozen or embedded in OCT were analyzed using targeted and non-targeted proteomics. A full description of methods used for targeted and non-targeted proteomics is also provided within the supplementary materials.

### Spatial RNA sequencing

Formalin-fixed paraffin-embedded (FFPE) tissues were processed for spatial transcriptomic profiling. 5µm FFPE tissue sections were placed on a 10x Genomics (Pleasanton, CA) Xenium slide and incubated at 42C for 3 hours, prior to being deparaffinized and decrosslinked via immersion in xylene and ethanol and hydrated in water to remove residual ethanol. Once hydrated, tissues slides were decrosslinked. Tissue slides were then inserted into 10x Genomics Xenium slide cassettes. Decrosslinking buffer was added to the well created by the cassette and incubated. After incubation, buffer was removed, and the sample was washed. Tissue was then processed through Hybridization, Ligation & Amplification protocols. These protocols are described fully within the supplementary materials.

For analysis of each Xenium sample, centroid (location), segmentation (cell boundaries), and the counts matrix data were extracted. Cells were filtered to have more than five features and more than ten counts. These were normalized and run through the Banksy functions (Prabhakar lab^49^) with a lambda of 0.2 for the Banksy matrix computation and a resolution of 1.0 for maximum clusters. Gene expression was calculated as an average of counts for each gene in each cluster and then normalized by dividing each cluster’s average gene expression by the maximum average cluster expression for that gene. Hierarchical cluster heatmaps to aid with annotation were generated using clustergrammer (Maayan lab^50^), following normalization of raw gene counts using the logCPM method, before transformation using the Z-score method. Most probable annotations were then mapped back onto the clusters and used to color code visuals by cell type. UMAPs were made for each grade (IV and low), and spatial plots were generated for each sample. These create an informative representative of the original FFPE slide. Niches were made by facet-wrapping these spatial plots by cell type, allowing direct visualization of each layer (cell type). Differential expression analyses highlighted immune-tumor dynamics in pro-inflammatory and immunosuppressive pathways.

### Statistical analysis plan

Up to 10 patients were to be enrolled in the treatment arm of this study, while up to 20 patients served as controls. The sample size selected was chosen for practical considerations in the setting of a phase 0 study, to allow descriptive analysis of the changes of the biologic endpoints in the evolocumab treated (cases) and untreated patients (controls). For analysis of the level of evolocumab within the resected or biopsied tumor, this was to be compared with resected or biopsied control patients who were not treated with evolocumab via either a Wilcoxon rank sum test or a two-sample t-test, with adjustment for multiple comparisons. Assessment of the relationship between serum and intratumoral levels of drug were to be performed by using a scatterplot to graphically show the relationship between these two measures with spearman’s rank correlation used to assess association. Within this study, experimental results are presented as mean ± SD unless otherwise specified. Statistical tests for all studies were completed using GraphPad v.8.4.3 (Prism). Asterisks were appended to graphs to represent the significance level of any difference (*p≤0.05, **p≤0.01, ***p≤0.001, ****p≤0.0001, p>0.05 not significant). Comparisons between groups were performed using a two-tailed Mann-Whitney U test. Correlation analyses were performed using the Pearson correlation coefficient.

## Data Availability

Targeted proteomic data have been deposited to the ProteomeXchange Consortium (PXD053215) via the Panorama Public repository and is restricted while under review. Xenium spatial transcriptomics data has been uploaded to GEO (accession number: GSE289588) and is similarly restricted while under review. All demographic, AE, flow cytometry and IHC/IF data are available in the main text.

## General

We are grateful to all patients and their caregivers who enrolled in this study. We are also grateful to the Knox Martin Foundation and the Langford Cregger Foundation for their philanthropic support. We are also grateful to the team at the Duke Brain Tumor Biorepository for their assistance and co-ordination in tissue management and acquisition. We would also like to acknowledge the assistance of the Molecular Genomics Core at the Duke Molecular Physiology Institute, Duke University School of Medicine, for the generation of data for the manuscript.

## Author contributions

Conceptualization: KS, MK

Methodology: KS, MWF, MJV, MK

Investigation: KS, MWF, MJV, AMC, ELT, KMH, COR, EEB, KEB, WCM, DMA, AD, HSF, MOJ, AF, SK, JHS, APP, SGG, CYL

Pathology: GYL, REM

Neuroradiology: EC

Spatial RNA sequencing, AMC, KS

Writing – original draft: KS, MWF, PEF, MK

Writing – review & editing: KS, MWF, MJV, AMC, ELT, KMH, COR, EEB, DMA, AD, HSF, MOJ, AF, SK, JHS, APP, SGG, CYL, PEF, MK

Statistical analysis: KS, EDB, JEH

Supervision: PEF, MK

## List of Supplementary Materials

**Supplementary Methods**

**Fig. S1.** Mechanism of PCSK9i for increasing surface MHC-I

**Fig. S2.** Trial study schema

**Fig. S3.** Clustering heatmap for grade IV gliomas

**Fig. S4.** Clustering heatmap for grade II/III gliomas

**Fig. S5.** H&E staining for grade IV tumor with high evolocumab titers (ID: 1007) used for paired ST/IHC/IF

**Fig. S6.** Paired ST/IHC/IF for grade II tumor with low evolocumab titers (ID: 1008)

**Fig. S7.** H&E staining for grade II tumor with low evolocumab titers (ID: 1008)

**Table S1.** Demographics of consented participants

**Table S2.** Toxicity summary of all Adverse Events among participants treated with evolocumab

**Table S3.** Toxicity summary of Adverse Events among participants possibly, probably, or definitely related to evolocumab

**Table S4.** Full inclusion & exclusion criteria for study

**Supplementary References**

