## Supplementary Methods, Figures & Tables for "A surgical window of opportunity trial evaluating the effect of the PCSK9 inhibitor evolocumab on tumoral MHC-I expression and CD8^+^ infiltration in glioma"

### Supplementary Materials:

**List:**


- **Supplementary Methods**
- **Fig. S1.** Mechanism of PCSK9i for increasing surface MHC-I
- **Fig. S2.** Trial study schema
- **Fig. S3.** Clustering heatmap for grade IV gliomas
- **Fig. S4.** Clustering heatmap for grade II/III gliomas
- **Fig. S5.** H&E staining for grade IV tumor with high evolocumab titers (ID: 1007) used for paired ST/IHC/IF
- **Fig. S6.** Paired ST/IHC/IF for grade II tumor with low evolocumab titers (ID: 1008)
- **Fig. S7.** H&E staining for grade II tumor with low evolocumab titers (ID: 1008)
- **Table S1.** Demographics of consented participants
- **Table S2.** Toxicity summary of all Adverse Events among participants treated with evolocumab
- **Table S3.** Toxicity summary of Adverse Events among participants possibly, probably, or definitely related to evolocumab
- **Table S4.** Full inclusion & exclusion criteria for study
- **Supplementary References**

#### Supplementary Methods

##### Mass Spectroscopy

For targeted assay development, samples were diluted to 200 µL with 5% deoxycholate and 10 mM dithiothreitol (DTT) followed by heating at 80 °C for 20 min using a Thermomixer (Eppendorf). After cooling, alkylation was performed with 25 mM iodoacetamide in the dark for 30 min followed by the addition of 100 µg of TPCK-trypsin (Worthington) and incubation at 37 °C for 2 h. After quenching with 1.5% trifluoroacetic acid and filtering, digests were analyzed by a 20 min microflow LC-MS/MS using a Waters ACQUITY UPLC (1 x 100 mm or 1 x 150 mm ACQUITY Premier CSH column; direct injection; 100 µL/min flow rate; 3-28% MeCN-0.1% formic acid) interfaced to a Thermo Exploris 480. Neat Evo was analyzed by data-dependent acquisition, and database searching was performed with Skyline[1] using the MS Amanda search engine with Percolator post-search validation[2,3]. Peptide uniqueness was predicted using NCBI BLASTp and Nextprot peptide uniqueness checker[4]. Candidate proteotypic peptides were analyzed in the evolocumab-spiked plasma using parallel reaction monitoring as previously described. Data were analyzed in Skyline and have been uploaded to the ProteomeXchange consortium (details in data and materials availability sub-section).

For targeted and non-targeted proteomics, tissue was rinsed free with cold phosphate-buffered saline and homogenized by sonication or bead beating in a 10:1 volume per tissue weight of 5% (w/v) sodium dodecyl sulfate in 50 mM triethylammonium bicarbonate. Based on detergent-compatible Bradford assay, 20 µg of each sample was reduced and alkylated and digested with 2 or 10 µg Sequencing Grade Modified Trypsin (Promega) using an S-trap micro device (Protifi) and 47 °C for 1 h. 200 fmol of SpikeTides TQL peptides were added to the S-trap along with trypsin. After elution, peptides were lyophilized, reconstituted in 0.2 % formic acid, and a study pool QC (SPQC) sample was made by mixing equal amounts of all samples. Approximately 1 µg of each sample, along with replicates of a study pool QC (SPQC), were loaded onto Evotip Pure tips. Targeted proteomic analysis used an Evosep One LC interfaced to a Thermo Exploris 480 and analyzed using a 100 sample-per-day (100SPD) LC method and PRM as described above. During method development, 100 µg of select samples were digested with 10 µg trypsin, and 30 µg of digested were analyzed by microflow LC-MS/MS as with the whole blood. Data was analyzed in Skyline and normalized to stable isotope-labeled internal standard (at 10 fmol/µg) to derive femtomol per microgram values of evolocumab in tumor.

Non-targeted proteomics used an Evosep One LC interfaced to a Thermo Orbitrap Astral using a 60 sample-per-day (60SPD) LC method and data-independent acquisition (DIA) in the Astral analyzer[5,6]. MS/MS used 150 x 4 m/z windows from 380-980 m/z, and automatic gain control target of 500%, 6 ms ion transfer time and normalized collision energy of 28. Data was analyzed with DIA-NN 1.8.2 beta 27 in library-free mode[7]. Raw data was converted to .dia before processing. Default settings were used with trypsin specificity and up to 2 missed cleavage, and N-terminal acetylation as a variable modification, Identification and quantification used a 1% precursor and protein group false discovery rate. Data was further filtered to include protein groups with no missing data and %coefficient of variation <50 across four analyses of a QC pool.

##### Spatial RNA sequencing

Xenium samples were processed by first adding a panel containing 480 custom gene padlock probes enriched in genes relevant to cell type and cell state, including glioma, neuronal and immune cells to the tissue. Each circularizable DNA probe contains two regions that hybridize to target RNA and a third region that encodes a gene-specific barcode. The two ends of the probes bind the target RNA and are ligated to generate a circular DNA probe. Following ligation, the circularized probe is amplified, producing multiple copies of the gene-specific barcode for each target. Tissue slides were then stained using the 10x Genomics Multimodal Cell Segmentation Kit. During Multimodal staining, antibodies used for cell segmentation bind their antigens in an overnight incubation, followed by post-incubation washes to remove excess antibodies. The Multimodal Cell Segmentation kit stains for cell nuclei, membranes, and cell interior that are inputs for the 10x Genomics automated morphology-based cell segmentation analysis pipeline. Prepared tissue slides were then loaded for imaging on the Xenium Analyzer for spatial transcriptomic analysis. Fluorescently labeled oligos bind to the amplified DNA probes. Cyclical rounds of fluorescent probe hybridization, imaging, and removal generated optical patterns specific for each barcode, which were converted into a gene identity. Identified transcripts were then visualized using Xenium Explorer software.

**Fig. S1. Mechanism of PCSK9i for increasing surface MHC-I**

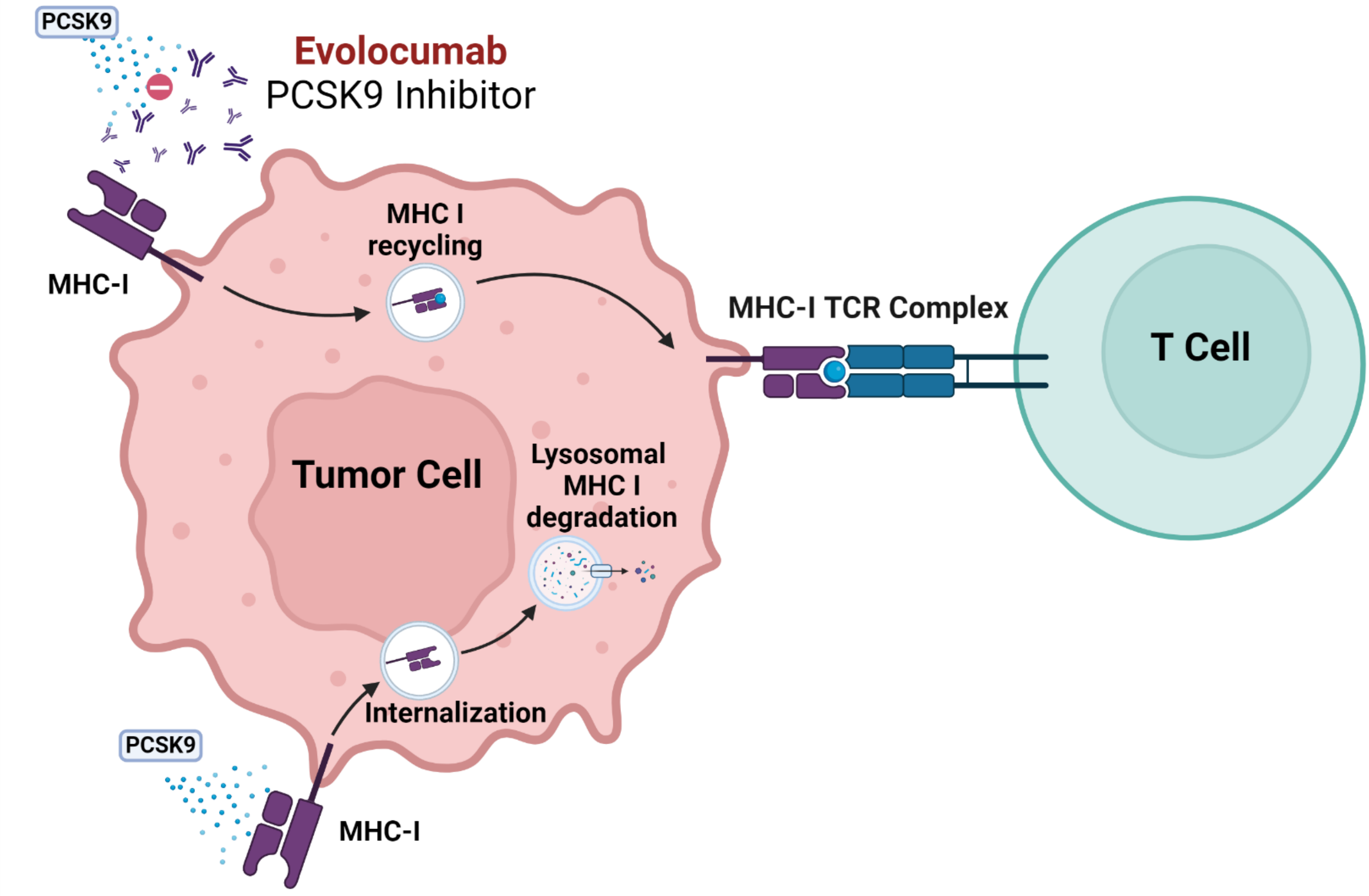


PCSK9 associates with MHC-I and promotes its degradation within intracellular lysosomes. Evolocumab inhibits PCSK9 by binding it and preventing it interacting with surface receptors such as MHC-I. MHC-I is instead recycled back to the cell surface, where it can present neoantigens to $CD8^{+}$ lymphocytes. Created with BioRender.com.

**Fig. S2. Trial study schema**

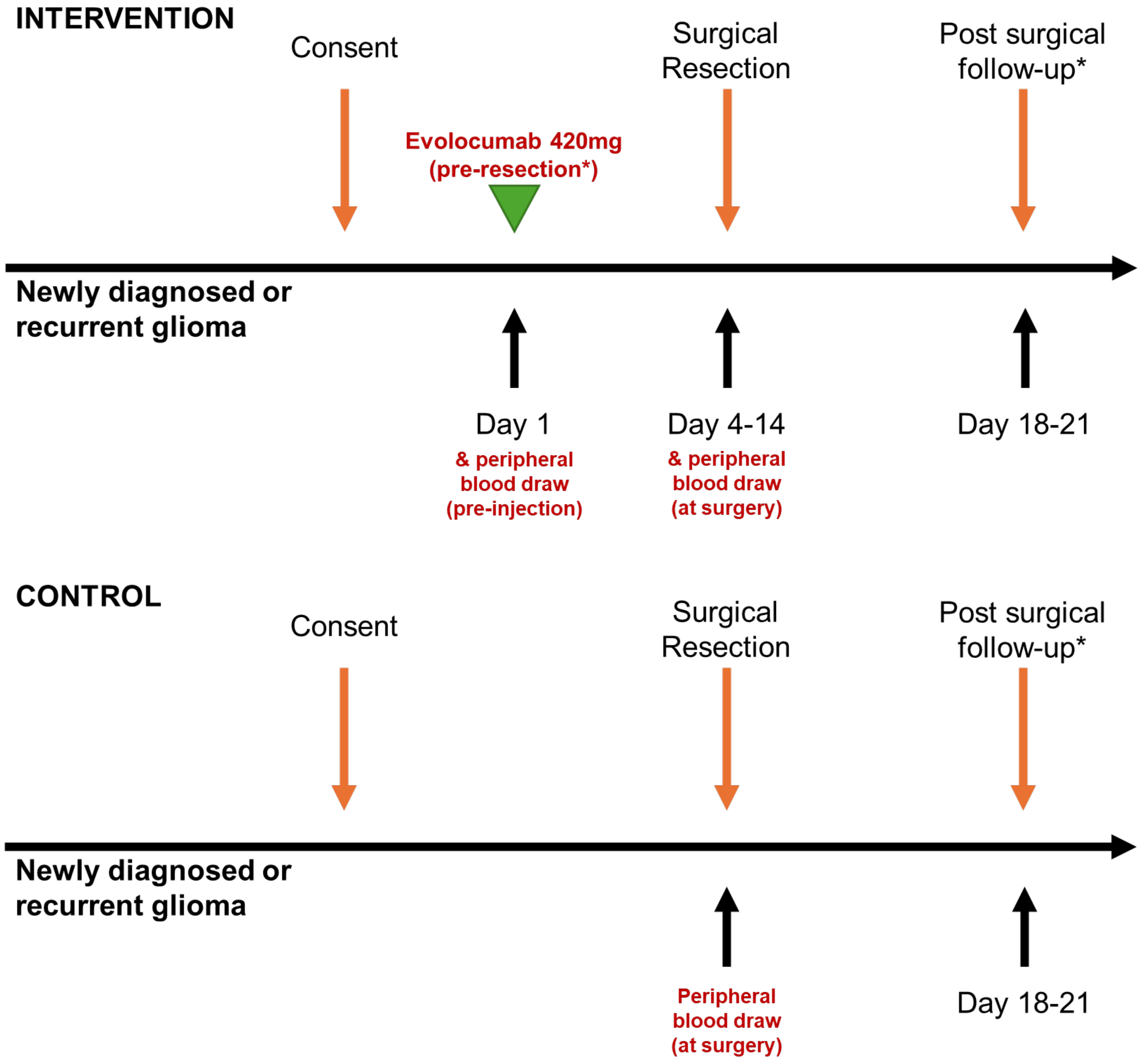

#### Fig. S3. Clustering heatmap for grade IV gliomas

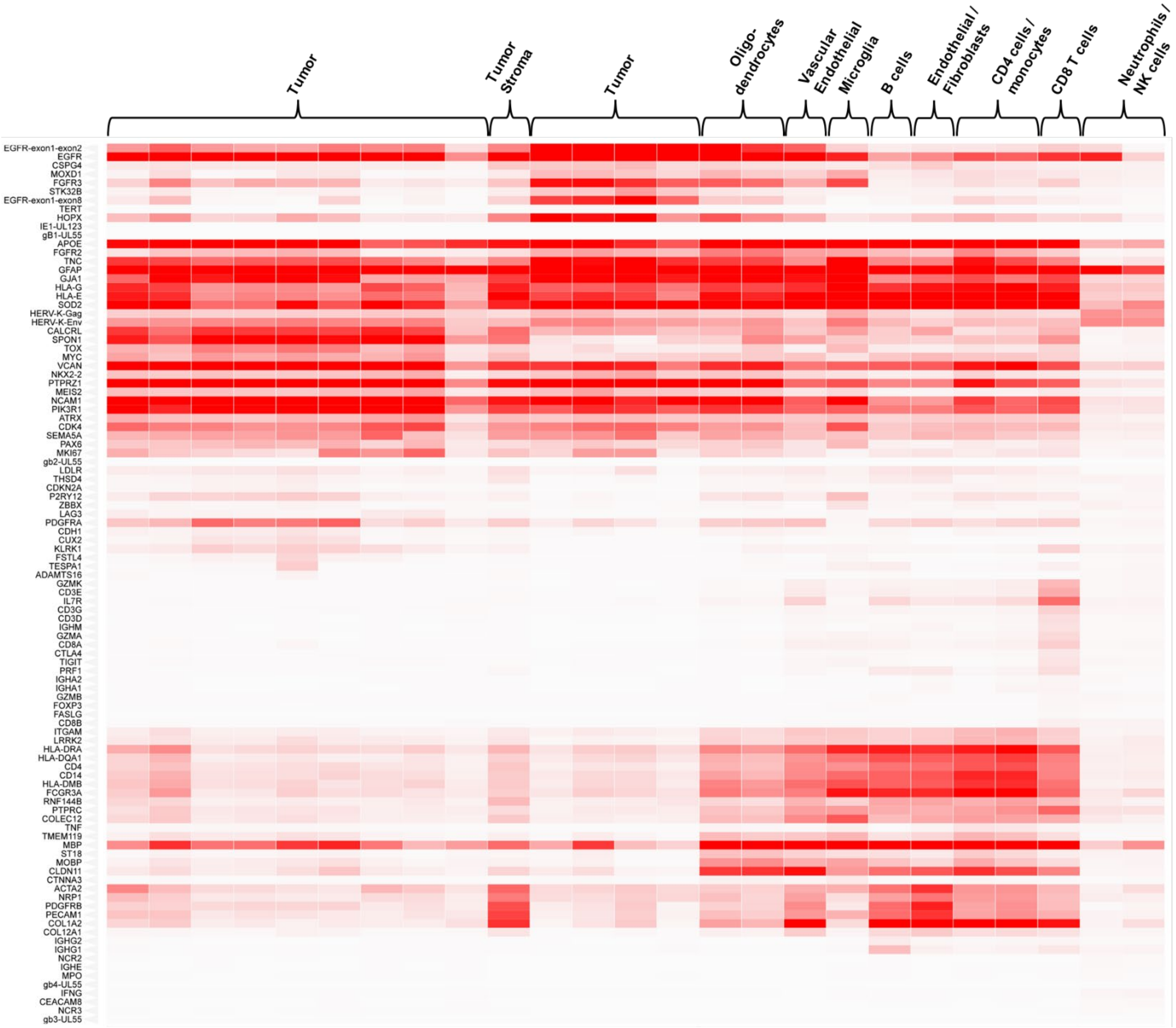


Hierarchical cluster heatmaps of gene transcripts from grade IV glioma samples shown, identifying regions of tumor, tumor stroma, oligodendrocytes, vascular endothelial cells, microglia, B cells, endothelial cells/fibroblasts, CD4 cells/monocytes, neutrophils/NK cells. Heatmaps generated using clustergrammer (Maayan Lab)

#### Fig. S4. Clustering heatmap for grade II/III gliomas

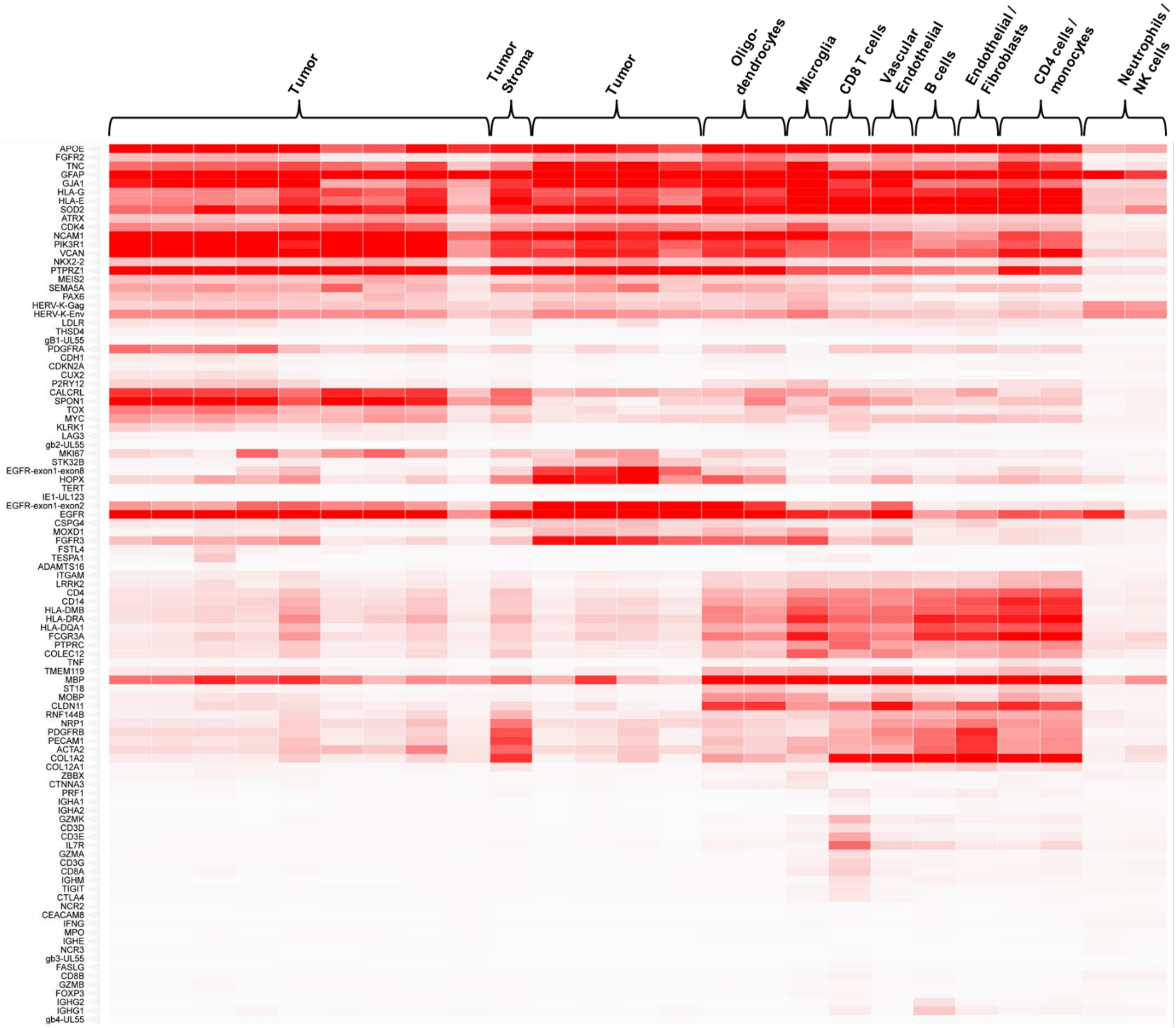


Hierarchical cluster heatmaps of gene transcripts from grade II/III glioma samples shown, identifying regions of tumor, tumor stroma, oligodendrocytes, microglia, CD8 T cells, vascular endothelial cells, B cells, endothelial cells/fibroblasts, CD4 cells/monocytes, neutrophils/NK cells. Heatmaps generated using clustergrammer (Maayan Lab)

**Fig. S5. H&E staining for grade IV tumor with high evolocumab titers (ID: 1007)**

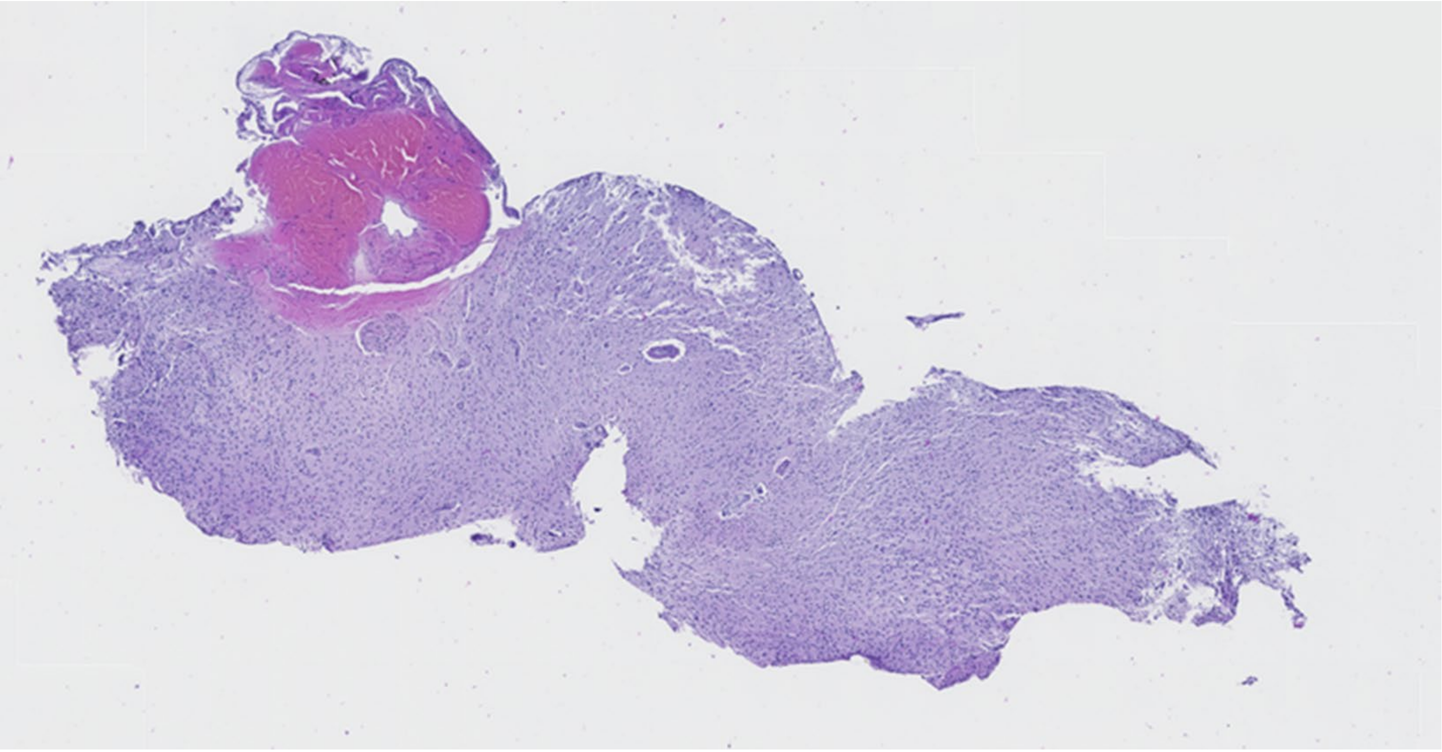

H&E sectioning of grade IV glioblastoma with high evolocumab titers used in paired spatial RNA sequencing/IHC/IF staining (ID: 1007, 20x magnification shown)

**Fig. S6. Paired spatial RNAseq/IHC/IF for grade II tumor with low evolocumab titers (ID: 1008)**

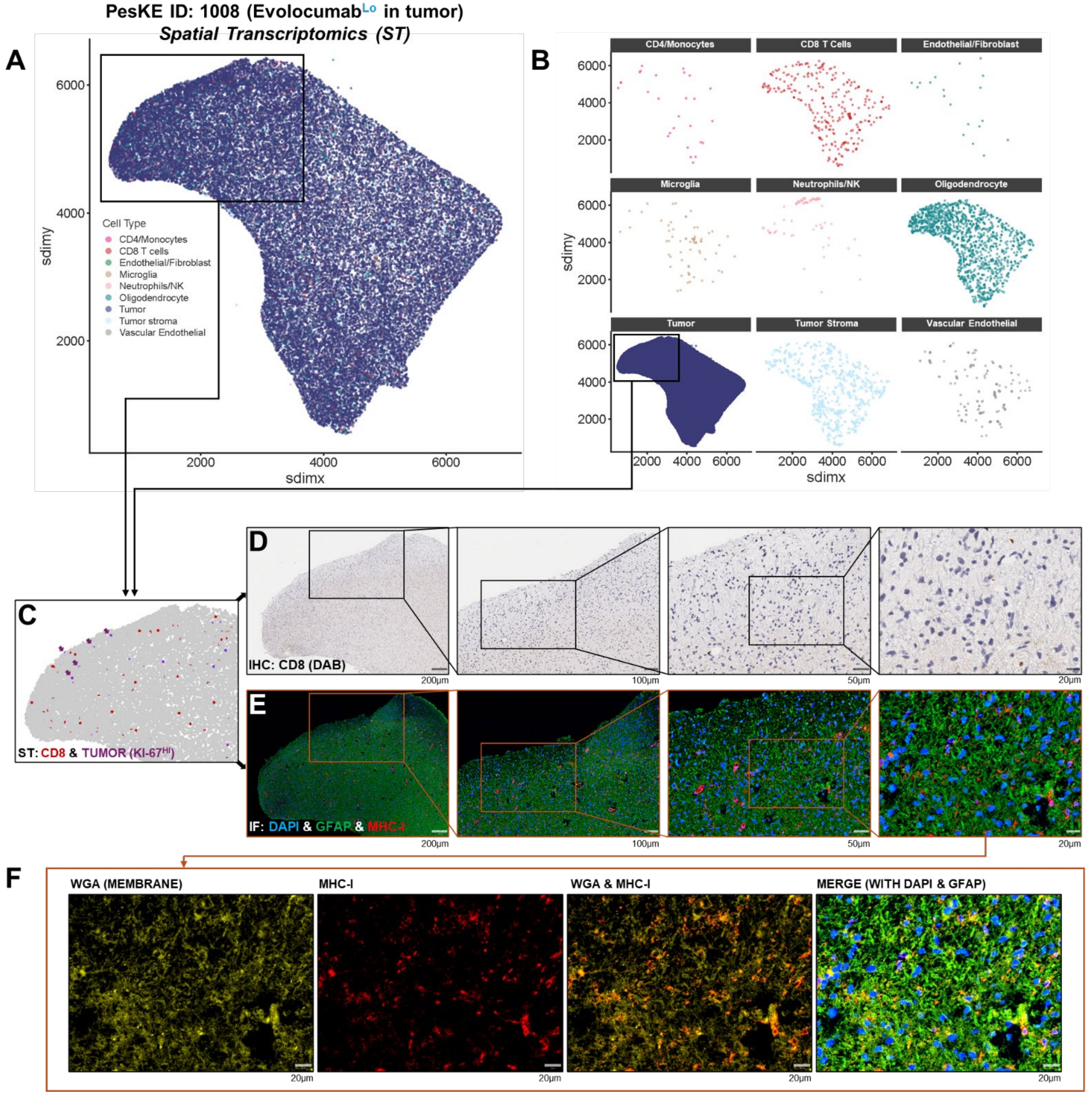


(A-C) Paired spatial RNA sequencing via Xenium platform and individual niches shown. Purple arrows in (C) identify proliferating tumor cells (Ki-67$^{Hi}$), overlapping with a denser region of tumor assessed via H&E. CD8$^{+}$ T cells are highlighted in red.

(C-E) Paired Immunohistochemistry (IHC) and Immunofluorescence (IF) from tissue samples from a case with low evolocumab titers (ID: 1008, grade II astrocytoma). Low levels of CD8$^{+}$ infiltration within the parenchyma are observed on IHC. IF staining of GFAP (green), DAPI (nuclei, blue) and MHC-I (red) shows no obvious increase in MHC-I expression or co-localization with cell membranes (shown via WGA (yellow)).

**Fig. S7. H&E staining for grade II tumor with low evolocumab titers (ID: 1008)**

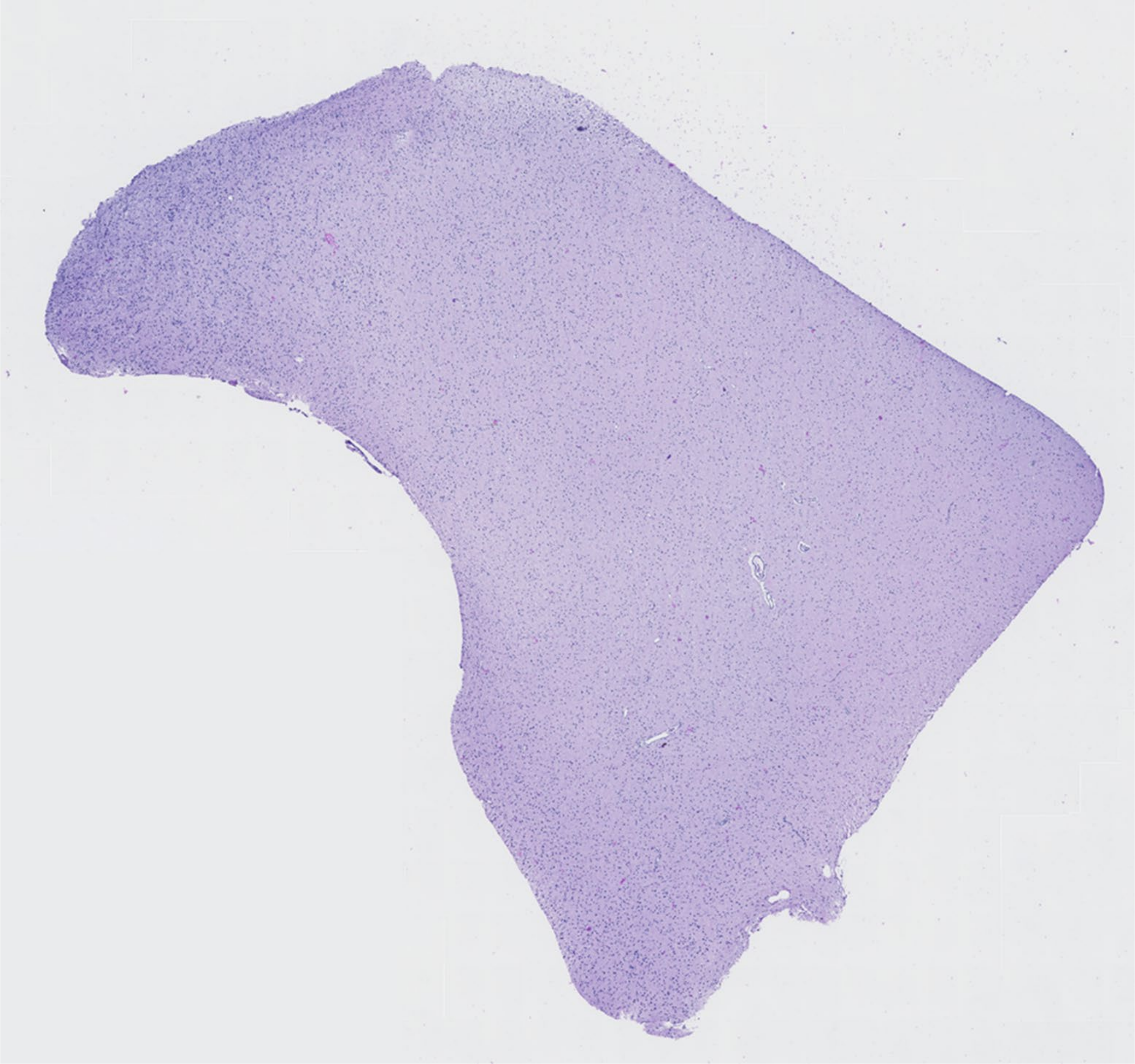

H&E sectioning of grade II astrocytoma with low evolocumab titers used in paired spatial RNA sequencing/IHC/IF staining (ID: 1008). The greatest intensity of staining is in the upper left-hand portion of tissue and migrates along the left-hand margin, typical of Grade 2 astrocytoma. 20x magnification shown.

**Table S1. Demographics of consented participants**

| Age at Consent | N | Mean | Std Dev | Min | Median | Max |
|---|---|---|---|---|---|---|
| Control | 26 | 51.85 | 16.07 | 28.00 | 49.00 | 79.00 |
| Evolocumab | 6 | 53.00 | 19.88 | 28.00 | 52.00 | 74.00 |

| | Study Group | | | | Total | |
|---|---|---|---|---|---|---|
| | Control | | Evolocumab | | | |
| | N | % | N | % | N | % |
| **Gender** | | | | | | |
| Male | 12 | 46.15 | 4 | 66.67 | 16 | 50.00 |
| Female | 14 | 53.85 | 2 | 33.33 | 16 | 50.00 |
| **Ethnicity** | | | | | | |
| Not Hispanic or Latino | 25 | 96.15 | 6 | 100.00 | 31 | 96.88 |
| Unknown | 1 | 3.85 | 0 | 0.00 | 1 | 3.13 |
| **Race** | | | | | | |
| White | 22 | 84.62 | 6 | 100.00 | 28 | 87.50 |
| Black or African American | 3 | 11.54 | 0 | 0.00 | 3 | 9.38 |
| Unknown | 1 | 3.85 | 0 | 0.00 | 1 | 3.13 |
| American Indian or Alaska Native | 0 | 0.00 | 0 | 0.00 | 0 | 0.00 |
| Asian | 0 | 0.00 | 0 | 0.00 | 0 | 0.00 |
| Native Hawaiian or other Pacific Islander | 0 | 0.00 | 0 | 0.00 | 0 | 0.00 |
| Not Reported | 0 | 0.00 | 0 | 0.00 | 0 | 0.00 |
| Multi-Race | 0 | 0.00 | 0 | 0.00 | 0 | 0.00 |
| **Histologic Grade** | | | | | | |
| 4 | 18 | 69.23 | 2 | 33.33 | 20 | 62.50 |
| Not Applicable | 6 | 23.08 | 2 | 33.33 | 8 | 25.00 |
| No tumor tissue analysis | 0 | 0.00 | 2 | 33.33 | 2 | 6.25 |
| 3 | 2 | 7.69 | 0 | 0.00 | 2 | 6.25 |
| **Surgical Pathology Diagnosis of Analyzed Sample** | | | | | | |
| Glioblastoma | 17 | 65.38 | 2 | 33.33 | 19 | 59.38 |
| Oligodendroglioma | 3 | 11.54 | 1 | 16.67 | 4 | 12.50 |
| No tumor tissue analysis | 0 | 0.00 | 2 | 33.33 | 2 | 6.25 |
| High Grade Astrocytoma, NOS | 2 | 7.69 | 0 | 0.00 | 2 | 6.25 |
| Low grade astrocytoma, NOS | 0 | 0.00 | 1 | 16.67 | 1 | 3.13 |
| Fibrillary Astrocytoma | 1 | 3.85 | 0 | 0.00 | 1 | 3.13 |

| | | | | | | |
|---|---|---|---|---|---|---|
| Other, specify: High grade glioma | 1 | 3.85 | 0 | 0.00 | 1 | 3.13 |
| Anaplastic Astrocytoma | 1 | 3.85 | 0 | 0.00 | 1 | 3.13 |
| Diffuse Astrocytoma | 1 | 3.85 | 0 | 0.00 | 1 | 3.13 |
| **Disease Status** | | | | | | |
| Newly Diagnosed | 16 | 61.54 | 1 | 16.67 | 17 | 53.13 |
| Previously Treated with Recurrent Disease | 9 | 34.62 | 3 | 50.00 | 12 | 37.50 |
| No tumor tissue analysis | 0 | 0.00 | 2 | 33.33 | 2 | 6.25 |
| Low grade treated, transformed | 1 | 3.85 | 0 | 0.00 | 1 | 3.13 |
| **Disease Description** | | | | | | |
| Unifocal | 24 | 92.31 | 4 | 66.67 | 28 | 87.50 |
| No tumor tissue analysis | 0 | 0.00 | 2 | 33.33 | 2 | 6.25 |
| Multifocal | 1 | 3.85 | 0 | 0.00 | 1 | 3.13 |
| Unknown | 1 | 3.85 | 0 | 0.00 | 1 | 3.13 |
| **Site of Surgical Pathology Tumor** | | | | | | |
| Frontal lobe | 10 | 38.46 | 1 | 16.67 | 11 | 34.38 |
| Temporal lobe | 8 | 30.77 | 0 | 0.00 | 8 | 25.00 |
| Parietal lobe | 5 | 19.23 | 0 | 0.00 | 5 | 15.63 |
| No tumor tissue analysis | 0 | 0.00 | 2 | 33.33 | 2 | 6.25 |
| Frontal-Parietal | 1 | 3.85 | 1 | 16.67 | 2 | 6.25 |
| Temporal-Occipital | 0 | 0.00 | 1 | 16.67 | 1 | 3.13 |
| Frontal-Temporal | 0 | 0.00 | 1 | 16.67 | 1 | 3.13 |
| Insula | 1 | 3.85 | 0 | 0.00 | 1 | 3.13 |
| Parietal-Temporal | 1 | 3.85 | 0 | 0.00 | 1 | 3.13 |
| **Total** | **26** | **100.00** | **6** | **100.00** | **32** | **100.00** |

***NOTE**: Although 6 patients enrolled in the evolocumab group, only 4 had both tumor tissue and blood collected on study.

**Table S2. Toxicity summary of all Adverse Events among participants treated with evolocumab**

| | Grade of Adverse Event | | | | | | | | | | |
|---|---|---|---|---|---|---|---|---|---|---|---|
| | 1- Mild | | 2- Mod | | 3-Severe | | 4-LifeThr | | 5-Lethal | | Treated Total |
| | n | (%) | n | (%) | n | (%) | n | (%) | n | (%) | N |
| **Non-Hematologic Adverse Events** | | | | | | | | | | | |
| GASTROINTESTINAL DISORDERS | | | | | | | | | | | |
| Gastritis | 0 | *(0%)* | 1 | *(17%)* | 0 | *(0%)* | 0 | *(0%)* | 0 | *(0%)* | 6 |
| GENERAL DISORDERS AND ADMINISTRATION SITE CONDITIONS | | | | | | | | | | | |
| Injection site reaction | 2 | *(33%)* | 0 | *(0%)* | 0 | *(0%)* | 0 | *(0%)* | 0 | *(0%)* | 6 |
| INFECTIONS AND INFESTATIONS | | | | | | | | | | | |
| Thrush | 1 | *(17%)* | 0 | *(0%)* | 0 | *(0%)* | 0 | *(0%)* | 0 | *(0%)* | 6 |
| METABOLISM AND NUTRITION DISORDERS | | | | | | | | | | | |
| Dehydration | 1 | *(17%)* | 0 | *(0%)* | 0 | *(0%)* | 0 | *(0%)* | 0 | *(0%)* | 6 |
| NERVOUS SYSTEM DISORDERS | | | | | | | | | | | |
| Seizure | 1 | *(17%)* | 0 | *(0%)* | 0 | *(0%)* | 0 | *(0%)* | 0 | *(0%)* | 6 |
| **Non-Hematologic Adverse Events** | | | | | | | | | | | |
| Summary | | | | | | | | | | | |
| **Maximum Non-Hematologic AE** | **4** | ***(67%)*** | **1** | ***(17%)*** | **0** | ***(0%)*** | **0** | ***(0%)*** | **0** | ***(0%)*** | **6** |
| **All Adverse Events** | | | | | | | | | | | |
| Summary | | | | | | | | | | | |
| **Maximum Overall AE** | **4** | ***(67%)*** | **1** | ***(17%)*** | **0** | ***(0%)*** | **0** | ***(0%)*** | **0** | ***(0%)*** | **6** |

**Table S3. Toxicity summary of Adverse Events among participants possibly, probably, or definitely related to evolocumab**

| | Grade of Adverse Event | | | | | | | | | | Treated Total |
|---|---|---|---|---|---|---|---|---|---|---|---|
| | 1- Mild | | 2- Mod | | 3-Severe | | 4-LifeThr | | 5-Lethal | | |
| | n | (%) | n | (%) | n | (%) | n | (%) | n | (%) | N |
| **Non-Hematologic Adverse Events** | | | | | | | | | | | |
| GENERAL DISORDERS AND ADMINISTRATION SITE CONDITIONS | | | | | | | | | | | |
| Injection site reaction | 2 | *(33%)* | 0 | *(0%)* | 0 | *(0%)* | 0 | *(0%)* | 0 | *(0%)* | 6 |
| **Non-Hematologic Adverse Events** | | | | | | | | | | | |
| Summary | | | | | | | | | | | |
| **Maximum Non-Hematologic AE** | **2** | ***(33%)*** | **0** | ***(0%)*** | **0** | ***(0%)*** | **0** | ***(0%)*** | **0** | ***(0%)*** | **6** |
| **All Adverse Events** | | | | | | | | | | | |
| Summary | | | | | | | | | | | |
| **Maximum Overall AE** | **2** | ***(33%)*** | **0** | ***(0%)*** | **0** | ***(0%)*** | **0** | ***(0%)*** | **0** | ***(0%)*** | **6** |

**Table S4. Full inclusion & exclusion criteria for study**

| Inclusion Criteria | Exclusion Criteria |
|---|---|
| **Both arms:** | **Treatment arm only:** |
| 1. Adult patients ≥ 18 years old | 1. Any patient with a history of a serious hypersensitivity reaction to evolocumab or any of the excipients in evolocumab |
| 2. Newly diagnosed glioma (diagnosis can be based on imaging) or recurrent glioma (if recurrent, prior pathology report demonstrating glioma is required) | 2. Patients with severe hepatic impairment outside of the range defined in the inclusion criteria within 7 days of starting evolocumab. |
| 3. A clinical indication for gross macroscopic resection, debulking of the glioma, or biopsy, with sufficient tumor size that can allow collection of specimens for the required analyses. | 3. History or evidence of central nervous system bleeding as defined by stroke or intraocular bleed (including embolic stroke) not associated with any antitumor surgery within 6 months before enrollment |
| **Treatment arm only:** | 4. Infection requiring intravenous antibiotics that was completed < 1 week of study enrollment (day 1) with the exemption of prophylactic antibiotics for long line insertion or biopsy |
| 4. Adequate hematologic function within 14 days prior to starting evolocumab defined as follows:<br>a. Hemoglobin ≥ 10 g/dL *(Note: the use of transfusion or other intervention to achieve Hgb ≥ 10.0 g/dl is acceptable)*<br>b. White Blood Cells ≥ 1.5 x $10^9$/L<br>c. Absolute Neutrophil Count (ANC) ≥ 1.0 x $10^9$/L<br>d. Platelets ≥ 100 x $10^9$/L or ≥ 50,000 for patients who received TMZ within the past year | 5. Females of reproductive potential and males who are unwilling to practice an acceptable method(s) of effective birth control while on study through 1 month (2 half-lives) after receiving the last dose of study drug. |
| 5. Adequate renal function within 14 days prior to starting evolocumab defined as calculated creatinine clearance (CrCL) of ≥ 30 mL/min/1.73$m^2$ by the Cockcroft-Gault formula | |
| 6. Adequate hepatic function within 14 days prior to starting evolocumab defined as follows:<br>a. Total bilirubin ≥ 1.5 x institutional upper limit of normal (ULN) *(Note: Patients with known Gilbert disease without other clinically significant liver abnormalities are not excluded.)*<br>b. AST(SGOT) and ALT(SGPT) ≥ 1.5 × ULN | |
| 7. Negative serum pregnancy test (in females of childbearing potential) within 48 hours of starting evolocumab. | |
